# Ventilatory Burden Predicts Change in Sleepiness Following Positive Airway Pressure in Sleep Apnea

**DOI:** 10.1101/2024.11.12.24316879

**Authors:** Eric Staykov, Dwayne L. Mann, Samu Kainulainen, Timo Leppänen, Juha Töyräs, Ali Azarbarzin, Scott A. Sands, Philip I. Terrill

## Abstract

**Rationale:** Excessive daytime sleepiness, an important symptom of obstructive sleep apnea (OSA), is commonly quantified using the Epworth Sleepiness Scale score (ESS). Baseline OSA severity measures (ventilatory burden, flow limitation, and hypoxemia) provide insights into OSA pathophysiology and could predict changes in sleepiness (i.e. change-in-ESS) following continuous positive airway pressure (CPAP) treatment.

**Objectives:** We hypothesized that change-in-ESS following CPAP treatment can be predicted from baseline polysomnography.

**Methods:** Associations between OSA severity measures and ESS were evaluated in 2332 participants, adjusting for age, sex, BMI, and total sleep time. Change-in-ESS prediction was evaluated using 213 CPAP treatment studies (HomePAP, BestAIR, and ABC) in three steps: severity measures were compared (adjusted regression, *n*=64), a prediction model was developed using baseline ventilatory burden and baseline ESS (*n*=139), and then evaluated in holdout participants (*n*=74).

**Measurements and Main Results:** In cross-sectional analysis, ESS was associated with ventilatory burden (0.45 points/SD; 95% CI 0.23−0.67), hypoxic burden (0.39; 0.17−0.62), the apnea-hypopnea index (AHI) (0.36; 0.14−0.59), and flow limitation severity (0.22; 0.01−0.43). Comparison analysis revealed that change-in-ESS was most strongly associated with baseline ventilatory burden (-1.08 points/SD; -2.13 to -0.05) and baseline ESS (-2.75; -3.83 to -1.69); the AHI association was weaker (-0.97; -2.01−0.05). Predicted change-in-ESS and actual change-in-ESS were correlated in holdout participants (adjusted *R²*=0.313); median [IQR] actual change-in-ESS of predicted responders (≥2-point ESS improvement, *n*=54, 73.0%) was -5.0 [-10.0 to -2.0] and non-responders was 0.0 [-1.0−1.0] (*P*<0.001).

**Conclusions:** Baseline ventilatory burden and baseline ESS were independently associated with change-in-ESS and could be used together to inform clinicians whether CPAP treatment will likely improve a patient’s sleepiness.

## Introduction

Worldwide, almost 425 million adults aged 30 to 69 years have moderate to severe obstructive sleep apnea (OSA), for which treatment is recommended (1). Excessive daytime sleepiness (hypersomnia) is an important symptom of OSA and is associated with adverse consequences such as increased risk of driving accidents (2). The Epworth Sleepiness Scale score (ESS) is the most common quantification of sleepiness in OSA, and is measured from 0 to 24 points using 8 questions on a 0- to 3-point Likert scale (3). Continuous positive airway pressure (CPAP) is the first-line treatment for OSA. Meta-analyses report that CPAP improves ESS by an average of 2.4–2.9 points more than placebo (4, 5). However, 28% of all treated patients and 19% of optimally treated patients (≥7 hours/night CPAP use) have residual sleepiness (ESS>10) (6, 7). Enabling clinicians to accurately predict a patient’s change-in-ESS following CPAP treatment using baseline data may improve CPAP prescription and patient uptake decisions.

Several studies have examined the relationship between baseline polysomnographic characteristics and change in sleepiness following CPAP treatment, with conflicting results. Bhat *et al.* found significant correlations between change-in-ESS and baseline apnea-hypopnea index (AHI) and baseline time below 90% oxygen saturation (T90) in severe OSA (8). However, Kingshott *et al.* reported no significant correlations between change-in-ESS and baseline AHI, arousal index, oxygen desaturation index (ODI), or minimum oxygen saturation in severe OSA (9). Another study showed that baseline AHI was not a predictor of change-in-ESS, but paradoxically lower baseline AHI was associated with greater improvement in the Pittsburgh Sleep Quality Index (PSQI) (10). Balakrishnan *et al.* demonstrated that baseline OSA severity category (mild, moderate, and severe) and a composite metric named the Sleep Apnea Severity Index (SASI) were both associated with CPAP-related change in PSQI (11). Several algorithms have recently been developed that quantify different domains of OSA pathophysiology from diagnostic polysomnography (Figure 1), i.e. ventilatory burden (12, 13), flow limitation severity and frequency (14, 15), and hypoxemia severity (16, 17). These measures have demonstrated stronger associations with outcomes such as sleepiness, vigilance, cardiovascular disease, and all-cause mortality than conventional clinical metrics such as the AHI (18–20). To date, the potential to predict CPAP-related change-in-ESS using advanced measures from baseline polysomnography has not been evaluated.

**Figure 1.**
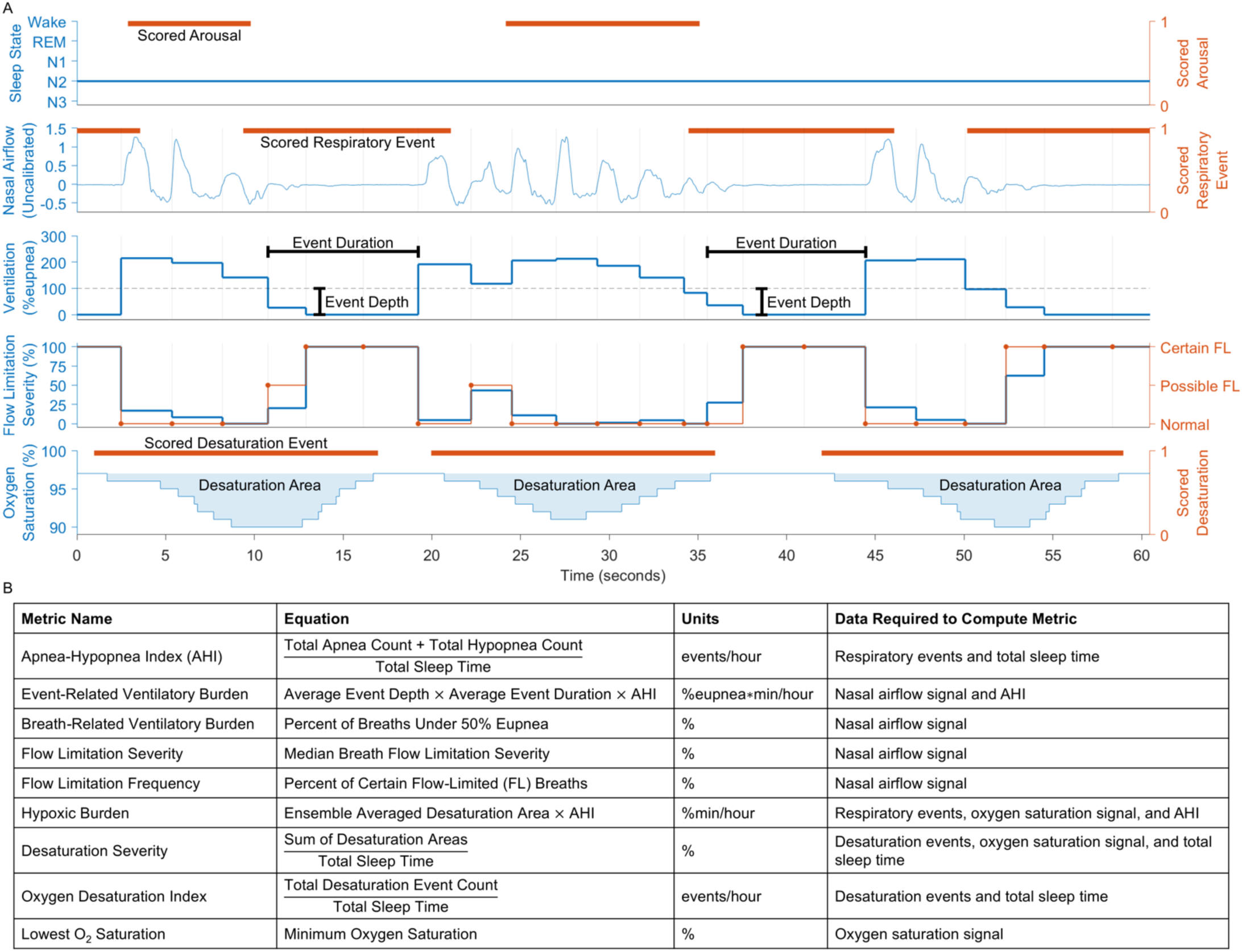
(*A*) Demonstration of the polysomnographic signals and scored events needed to compute the metrics used in this study, e.g. ventilatory burden, airway flow limitation, and hypoxemia severity. The red horizontal bars represent manually scored arousal, respiratory, or desaturation events. Ventilation and flow limitation were quantified on a breath-by-breath basis, with vertical gray lines denoting the start of automatically detected breaths. “Event-related ventilatory burden” is cumulative lost ventilation per hour during sleep using 90% eupnea as an indication of hypoventilation (12). “Breath-related ventilatory burden” is the percentage of breaths during sleep under 50% eupnea, modified from Parekh *et al.* (13). Where possible, only breaths during sleep excluding arousals were used to compute breath-related metrics (i.e. breath-related ventilatory burden, flow limitation severity, and flow limitation frequency). Airflow shapes were used to estimate flow limitation using continuous (flow limitation severity) and discrete (flow limitation certainty) scales (14, 15). (*B*) Table of equations, units, and data required to compute the metrics. FL = flow-limited.

The overarching goal of this study was to determine whether baseline polysomnographic characteristics can predict change-in-ESS following CPAP treatment. We hypothesized that: 1) physiology-based measures of OSA severity (ventilatory burden, flow limitation, and hypoxemia severity) would be associated with both baseline ESS and change-in-ESS, independent of confounders and mediators; 2) change-in-ESS can be predicted from baseline polysomnography, and that physiology-based measures of OSA severity would have more predictive ability than conventional measures of OSA severity such as the AHI; and 3) predicted change-in-ESS can be used to stratify responders (≥2-point ESS improvement (21, 22)) from non-responders. Finally, we sought to provide a chart that allows clinicians to use baseline polysomnography and patient characteristics to predict change-in-ESS following CPAP treatment.

## Methods

Below are key methodological details. More specific details are provided in the Supplementary Methods. Figure 2 shows a flow chart of the data analysis steps in this study.

**Figure 2.**
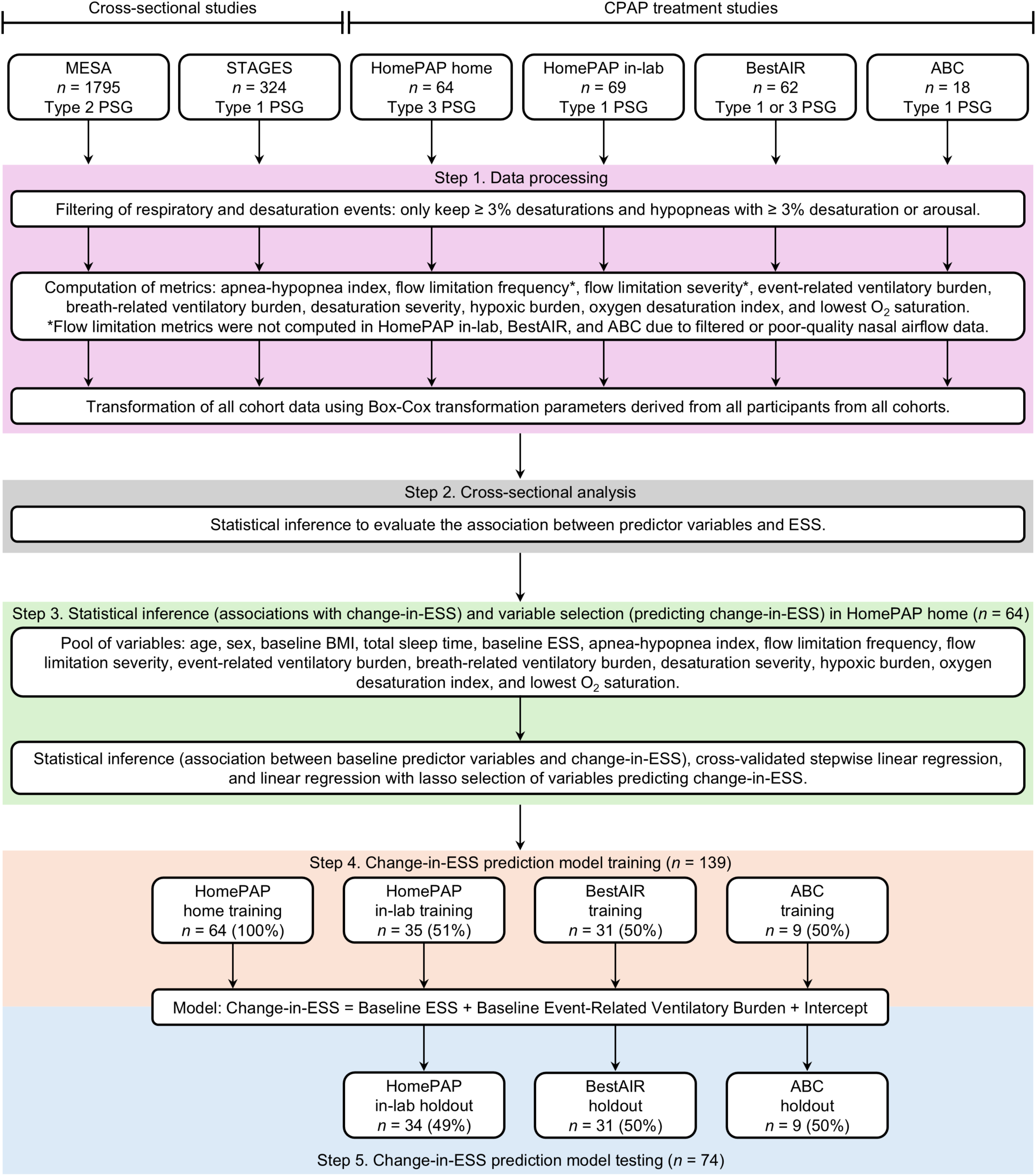
Flow chart of the data analysis steps in this study. Note that no HomePAP home participants were included in the holdout dataset because all 64 participants were used for statistical inference and variable selection (Step 3). PSG = polysomnography; ESS = Epworth Sleepiness Scale score.

### Study design and participants

This study used data from MESA (*n*=1795), STAGES (*n*=324), HomePAP (*n*=133), BestAIR (*n*=62), and ABC (*n*=18) studies. MESA is an ethnically diverse community sample of participants without known cardiovascular disease. The STAGES cohort consists of participants with suspected OSA who underwent diagnostic polysomnography. In MESA and STAGES, participants had a single ESS measurement and polysomnography. HomePAP, BestAIR, and ABC participants had a baseline ESS measurement and diagnostic polysomnography prior to CPAP treatment, and a follow-up ESS measurement after the treatment period. The treatment period varied between participants, with the median [IQR] being 3 [3–6] months. HomePAP participants had baseline AHI≥15 events/hour and ESS≥12 points and were randomized to home or in-lab diagnostic/management pathways. The BestAIR study recruited participants with prior cardiovascular disease who were not severely sleepy (ESS≤14 points) to examine the effect of CPAP treatment on blood pressure. ABC participants had at least one OSA symptom, BMI between 35−45 kg/m^2^, and AHI ≥20 events/hour; and were randomized to CPAP and bariatric surgery groups. Individual study details are reported in respective publications (19, 20, 23–25). Participant exclusion criteria are presented in Figure E1. Analyses were approved by The University of Queensland Research Ethics and Integrity unit (2023/HE000064, 2023/HE001953, and 2021/HE002256).

### Quantifications of OSA severity

For each cohort, consistent respiratory and desaturation event criteria were applied before computing OSA severity metrics. OSA severity was quantified using two measures of ventilatory burden (12, 13), two measures of flow limitation (14, 15), two measures of hypoxemia severity (16, 17), the SASI (26), and conventional clinical metrics. “Event-related ventilatory burden” is cumulative lost ventilation per hour during sleep (12), and “breath-related ventilatory burden” is the percentage of breaths during sleep under 50% eupnea, modified from Parekh *et al.* (13).

### Cross-sectional and CPAP treatment study analyses

All continuous variables were transformed for normality and standardized (Table E1). To determine which predictor variables are associated with baseline sleepiness and might therefore be predictive of change-in-ESS, associations between predictor variables and ESS were evaluated in 2332 participants across MESA and STAGES cohorts and HomePAP, BestAIR, and ABC studies (using their baseline ESS and diagnostic polysomnography prior to CPAP treatment). Multivariable linear regression models were adjusted for age, sex, BMI, and total sleep time. The significance level threshold was *P*<0.05.

Change-in-ESS prediction was evaluated in 213 participants across HomePAP, BestAIR, and ABC studies in three steps. First, relationships between baseline OSA severity measures and change-in-ESS were evaluated using multivariable linear regression in the HomePAP home cohort, noting that the polysomnograms from these studies had unfiltered and high signal-to-noise ratio nasal airflow which is needed to confidently compute flow limitation metrics. Linear regression models were adjusted for: 1) baseline ESS; 2) age, sex, and baseline BMI; and 3) age, sex, baseline BMI, total sleep time, baseline ESS, and nightly CPAP use. Next, cross-validated stepwise linear regression was performed using HomePAP home studies to determine the best combination of predictors of change-in-ESS, which were further confirmed using an alternative approach known as LASSO regression. Finally, data from HomePAP, BestAIR, and ABC studies (*n*=139) were used to train linear regression models to predict change-in-ESS using baseline OSA severity measures. These models were tested in holdout participants from these cohorts (*n*=74). Participants were ordered by their identification number and alternatingly allocated to training and holdout datasets.

## Results

Participant characteristics are summarized in Tables 1 and 2. Histograms showing distributions of variables and scatter plots showing relationships between variables for all participants from all cohorts are presented in Figure E2. In the combined cohort consisting of 2332 participants (Figure 3), increased ESS was associated with increased event-related ventilatory burden (0.45 points/SD; 95% CI 0.23−0.67), hypoxic burden (0.39; 0.17−0.62), AHI (0.36; 0.14−0.59), and flow limitation severity (0.22; 0.01−0.43).

**Figure 3.**
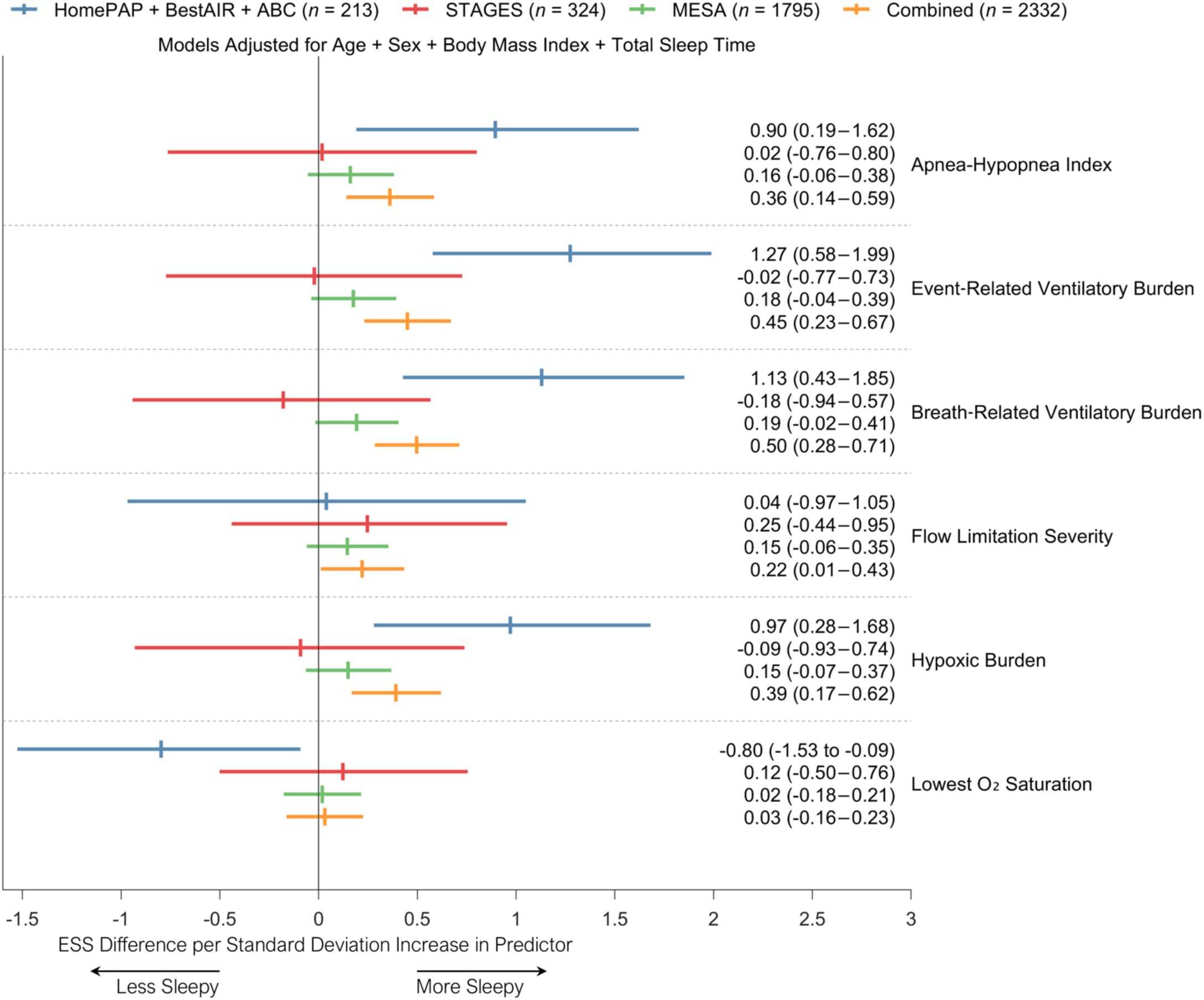
Association between 1-SD increase in predictor variables and Epworth Sleepiness Scale score (ESS) in cross-sectional cohorts (STAGES and MESA) and HomePAP, BestAIR, and ABC studies (using their baseline ESS and diagnostic polysomnography prior to continuous positive airway pressure treatment). Vertical ticks represent coefficient estimates and horizontal bars represent 95% confidence intervals. The combined cohort (*n* = 2332, orange bars) was composed of *n* = 213 participants from HomePAP, BestAIR, and ABC studies (blue bars); *n* = 324 participants from the STAGES cohort (red bars); and *n* = 1795 participants from the MESA cohort (green bars). Multivariable linear regression models were adjusted for age, sex, BMI, and total sleep time. Flow limitation frequency, desaturation severity, and oxygen saturation index could not be quantified in the STAGES cohort and are not present in this analysis. In HomePAP, BestAIR, and ABC studies, flow limitation severity could only be quantified in the HomePAP home studies (*n* = 64). Hypoxic burden could not be quantified in four MESA participants. One HomePAP in-lab participant and three MESA participants were excluded due to missing BMI data. One STAGES participant was excluded due to missing ESS data.

**Table 1.** Comparison of demographic, polysomnographic, and Epworth Sleepiness Scale score (ESS) characteristics of the STAGES and MESA cohorts.

|  | **STAGES**<br>**Cohort** | **MESA**<br>**Cohort** |
| --- | --- | --- |
| **Participants *n* (% male)** | 324 (54.0%) | 1795 (46.7%) |
| **Age (years)** | 42.0 [30.5–56.5] | 67.0 [61.0–75.0] |
| **Body Mass Index (kg/m<sup>2</sup>)** | 27.2 [23.1–32.0] | 27.9 [24.7–31.7] |
| **Depression (dichotomous)** | 56 (17.3%) | 304 (16.9%) |
| **Sleep Apnea Severity Index** |  |  |
| **Mild (I)** | 252 (77.8%) | 1498 (83.5%) |
| **Moderate (II)** | 60 (18.5%) | 198 (11.0%) |
| **Severe (III)** | 11 (3.4%) | 96 (5.3%) |
| **Total Sleep Time (hours)** | 6.4 [4.1–7.6] | 6.2 [5.3–7.0] |
| **Apnea-Hypopnea Index (events/hour)** | 10.3 [4.9–18.3] | 19.7 [10.9–34.4] |
| **Nasal Airflow Signal-to-Noise Ratio (dB)** | 31.4 [29.0–33.4] | 37.7 [35.2–40.1] |
| **Flow Limitation Frequency (%)<sup>†</sup>** | Not computed | 7.9 [2.9–19.3] |
| **Flow Limitation Severity (%)** | 27.8 [19.9–36.8] | 31.9 [24.9–39.2] |
| **Event-Related Ventilatory Burden (%eupnea\*min/hour)** | 89.0 [37.8–213.9] | 261.5 [116.4–566.0] |
| **Breath-Related Ventilatory Burden (%)** | 3.4 [1.9–7.0] | 5.8 [2.7–12.8] |
| **Desaturation Severity (%)<sup>‡</sup>** | Not computed | 0.8 [0.4–1.6] |
| **Hypoxic Burden (%min/hour)** | 11.8 [4.5–30.6] | 39.2 [20.9–74.8] |
| **Oxygen Desaturation Index (events/hour)<sup>‡</sup>** | Not computed | 24.5 [13.9–42.6] |
| **Lowest Oxygen Saturation (%)** | 83.0 [78.0–88.0] | 76.0 [67.0–81.0] |
| **ESS (points)** | 8.0 [5.0–12.0] | 5.0 [3.0–8.0] |

Data are presented as *n* (%) for categorical variables or median [IQR] for untransformed variables.
<sup>†</sup>Flow limitation frequency was not computed using STAGES polysomnograms which had relatively low nasal airflow signal-to-noise ratio and no way of correcting for this like flow limitation severity.
<sup>‡</sup>Desaturation severity and oxygen desaturation index were not computed using STAGES polysomnograms because desaturation scoring was unavailable.

**Table 2.** Comparison of demographic, polysomnographic, and Epworth Sleepiness Scale score (ESS) characteristics of the HomePAP home, HomePAP in-lab, BestAIR, and ABC cohorts.

|  | HomePAP Home Cohort | HomePAP In-Lab Cohort | BestAIR Cohort | ABC Cohort |
| --- | --- | --- | --- | --- |
| **Participants *n* (% male)** | 64 (67.2%) | 69 (53.6%) | 62 (71.0%) | 18 (55.6%) |
| **Age (years)** | 50.0 [42.5–58.5] | 47.0 [39.0–55.2] | 66.5 [60.0–70.0] | 48.8 [40.4–55.6] |
| **Baseline Body Mass Index (kg/m<sup>2</sup>)** | 37.0 [31.7–43.5] | 35.7 [32.1–45.0] | 31.2 [26.3–34.5] | 39.0 [35.8–40.4] |
| **Follow-up Body Mass Index (kg/m<sup>2</sup>)** | 37.2 [31.9–43.3] | 35.8 [32.4–44.7] | 31.6 [26.6–35.0] | 36.5 [35.0–39.8] |
| **Depression (dichotomous)** | 18 (28.1%) | 17 (24.6%) | 17 (27.4%) | No data |
| **Sleep Apnea Severity Index** |  |  |  |  |
| **Mild (I)** | 11 (17.2%) | 9 (13.0%) | 42 (67.7%) | 7 (38.9%) |
| **Moderate (II)** | 19 (29.7%) | 36 (52.2%) | 17 (27.4%) | 6 (33.3%) |
| **Severe (III)** | 34 (53.1%) | 23 (33.3%) | 3 (4.8%) | 5 (27.8%) |
| **Total Sleep Time (hours)** | 5.7 [5.1–6.1] | 5.4 [3.2–6.4] | 6.7 [5.7–7.6] | 6.6 [5.8–7.2] |
| **Apnea-Hypopnea Index (events/hour)** | 43.7 [29.0–80.1] | 36.1 [22.9–53.8] | 32.2 [22.3–38.3] | 40.7 [30.6–54.1] |
| **Nasal Airflow Signal-to-Noise Ratio (dB)** | 41.5 [38.2–45.3] | 41.2 [36.8–45.4] | 29.4 [23.3–32.9] | 25.6 [20.5–27.7] |
| **Flow Limitation Frequency (%)<sup>†</sup>** | 39.2 [23.1–58.4] | Not computed | Not computed | Not computed |
| **Flow Limitation Severity (%)<sup>†</sup>** | 35.8 [26.6–49.3] | Not computed | Not computed | Not computed |
| **Event-Related Ventilatory Burden (%eupnea\*min/hour)** | 842.6 [446.4–1998.8] | 694.1 [357.2–934.7] | 462.5 [346.0–647.3] | 621.6 [380.0–907.7] |
| **Breath-Related Ventilatory Burden (%)** | 15.3 [9.7–39.5] | 18.2 [9.7–29.3] | 10.4 [6.7–14.1] | 17.5 [8.5–22.8] |
| **Desaturation Severity (%)** | 1.3 [0.7–2.4] | 1.6 [1.1–2.1] | 0.7 [0.5–1.1] | 1.2 [0.8–1.7] |
| **Hypoxic Burden (%min/hour)** | 129.0 [71.4–201.8] | 85.1 [56.9–113.8] | 72.2 [58.2–112.9] | 78.2 [66.6–115.2] |
| **Oxygen Desaturation Index (events/hour)** | 45.3 [34.1–83.0] | 48.3 [30.4–66.0] | 30.0 [25.1–38.1] | 47.3 [41.2–61.4] |
| **Lowest Oxygen Saturation (%)** | 71.5 [61.5–79.0] | 77.0 [70.0–82.0] | 79.5 [75.0–82.0] | 81.0 [74.0–82.0] |
| **Nightly CPAP Use (hours)** | 5.1 [3.2–6.3] | 3.0 [1.4–5.3] | No data | No data |
| **Baseline ESS (points)** | 14.0 [12.0–16.0] | 14.0 [12.0–17.2] | 8.0 [5.0–10.0] | 9.0 [7.0–14.0] |
| **Follow-up ESS (points)** | 7.0 [4.0–9.0] | 7.0 [4.0–11.0] | 6.0 [3.0–8.0] | 6.0 [4.0–11.0] |
| **Change-in-ESS (points)** | -8.0 [-10.0 to -4.0] | -6.0 [-10.0 to -2.0] | -1.5 [-4.0–0.0] | -2.0 [-6.0–0.0] |
| **Change-in-ESS (%)** | -53.3 [-67.4 to -34.8] | -47.1 [-72.9 to -19.2] | -14.3 [-50.0–0.0] | -21.9 [-50.0–0.0] |
| **Follow-up Duration (months)** | 3.0 [3.0–3.0] | 3.0 [3.0–3.0] | 6.0 [6.0–6.0] | 9.0 [9.0–9.0] |

Data are presented as *n* (%) for categorical variables or median [IQR] for untransformed variables.
<sup>†</sup>Flow limitation frequency and severity could only be quantified in HomePAP home polysomnograms which had unfiltered and high signal-to-noise ratio nasal airflow data.

Multivariable linear regression (Figures 4 and E3) revealed that change-in-ESS was most strongly associated with baseline event-related ventilatory burden (-1.08 points/SD; -2.13 to -0.06) and baseline ESS (-2.75; -3.83 to -1.69); the AHI association was weaker (-0.97; -2.01−0.05). Event-related ventilatory burden was significantly associated with change-in-ESS after adjusting for age, sex, and baseline BMI (-1.46; -2.78 to -0.17), and borderline significant after further adjusting for total sleep time, baseline ESS, and nightly CPAP use (-0.99; -2.15−0.15). Variable selection was performed in a subset of the training dataset (HomePAP home studies, *n*=64). Baseline event-related ventilatory burden and baseline ESS was the most frequently selected combination of predictors of change-in-ESS using cross-validated stepwise linear regression (Table E2). They were also the first and only two variables to be selected using cross-validated LASSO selection of variables (Figure E4).

**Figure 4.**
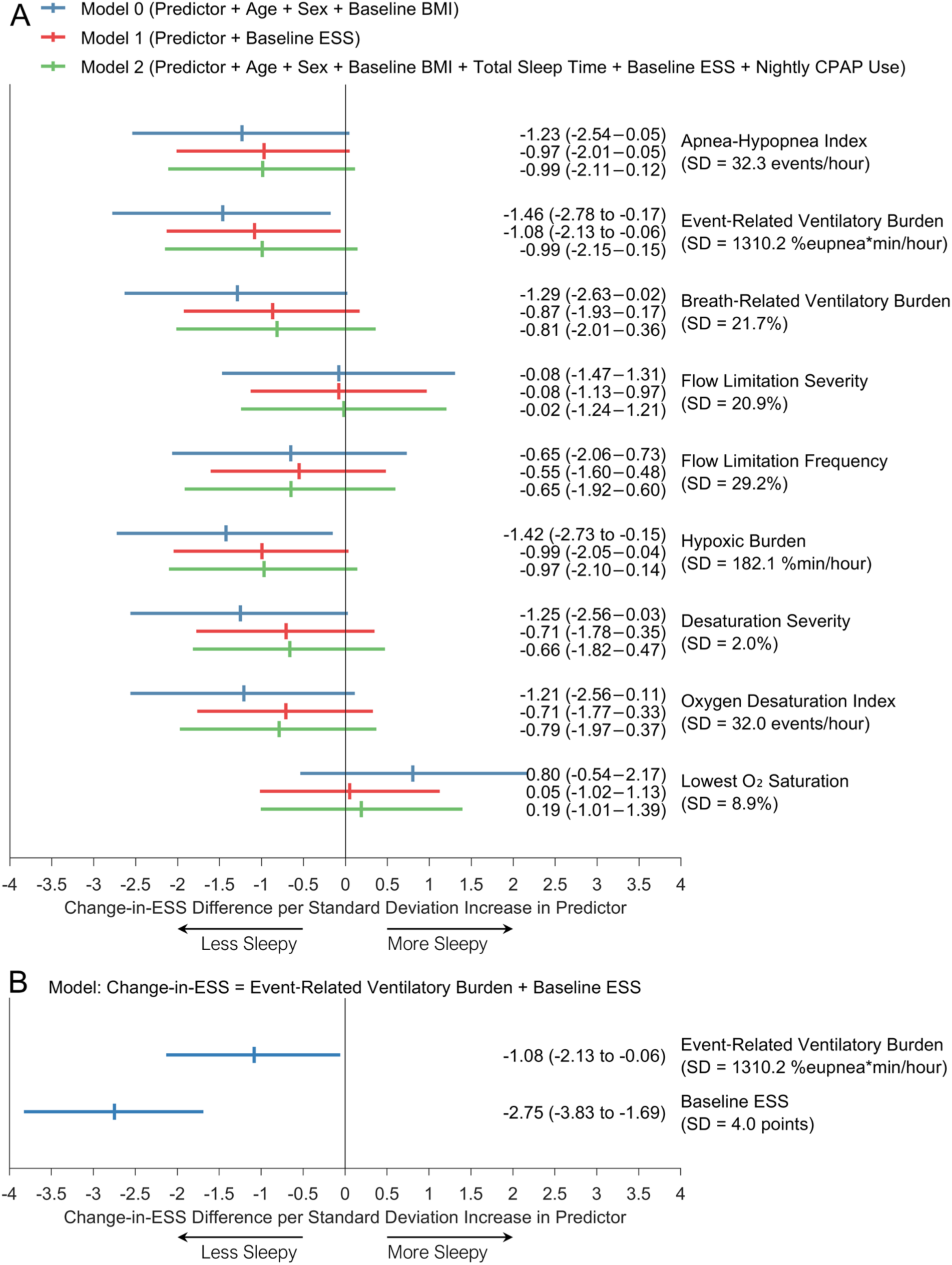
Association between 1-SD increase in baseline predictor variables and change-in-ESS in the HomePAP home polysomnography cohort. Vertical ticks represent coefficient estimates and horizontal bars represent 95% confidence intervals. (*A*) Linear regression models were adjusted for age, sex, and baseline BMI (model 0, blue bars); adjusted for baseline ESS (model 1, red bars); and further adjusted for age, sex, baseline BMI, total sleep time, and nightly CPAP use (model 2, green bars). (*B*) A linear regression model with baseline event-related ventilatory burden and baseline ESS. All analyses had *n* = 64 participants except for those adjusting for nightly CPAP use, which had *n* = 63 due to one participant missing this data. ESS = Epworth Sleepiness Scale score.

The linear regression model developed using both baseline event-related ventilatory burden and baseline ESS provided an adjusted *R²* (between predicted change-in-ESS and actual change-in-ESS) of 0.484 in the training dataset and 0.313 in the holdout dataset (Figure 5). Heteroscedasticity and bias were absent in Bland-Altman plots. In a comparative analysis, the use of baseline hypoxic burden rather than baseline event-related ventilatory burden provided an adjusted *R²* of 0.296 in the holdout dataset; the use of baseline AHI rather than baseline event-related ventilatory burden provided a value of 0.272; the use of baseline ESS alone provided a value of 0.234; and the use of the SASI provided a value of 0.228 (Table 3). The linear regression model provided a dichotomous prediction of ≥2-point ESS improvement (21, 22) after defining an optimal threshold (receiver operating characteristic curve, see Supplementary Figure E5). In the holdout dataset, predicted responders exhibited a median [IQR] change-in-ESS of -5.0 [-10.0 to -2.0], which was significantly different from predicted non-responders (0.0 [-1.0−1.0], *P*<0.001, Figure 6). The classifier had 91.5% sensitivity, 59.3% specificity, and 79.7% accuracy. In sensitivity analyses, similar performance was observed when the change-in-ESS responder/non-responder threshold was varied from -1 to -9 (Table E3). In a comparative analysis, the use of baseline hypoxic burden rather than baseline event-related ventilatory burden had an accuracy of 77.0% in the holdout dataset; the use of baseline AHI rather than baseline event-related ventilatory burden had an accuracy of 77.0%; the use of baseline ESS alone had an accuracy of 77.0%; and the use of the SASI had an accuracy of 62.2% (Table E4). Higher sensitivities, lower specificities, and lower accuracies were observed when false positives had half the penalty weight of false negatives (Figures E6−E7 and Tables E5−E6).

**Figure 5.**
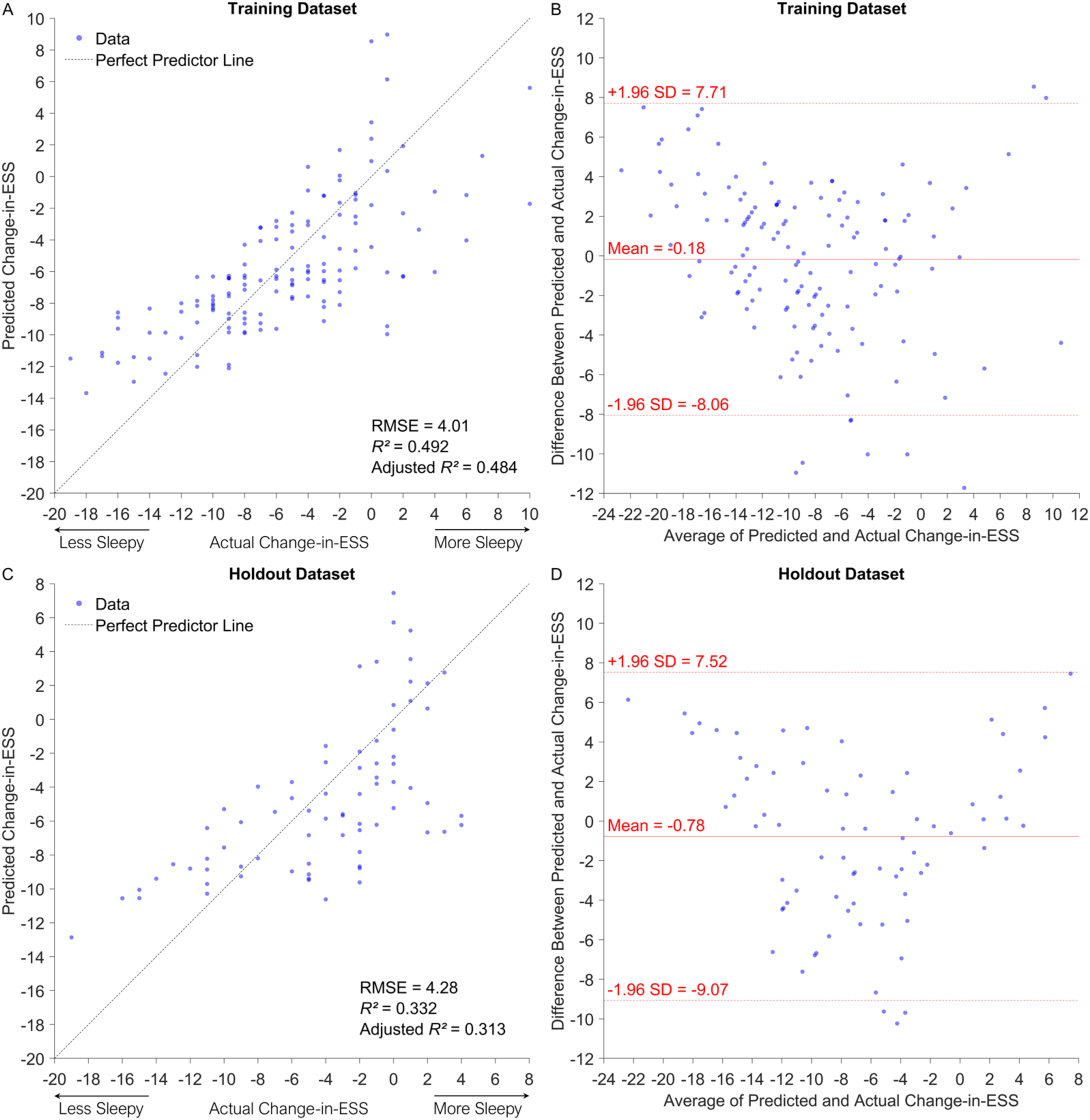
Linear regression model change-in-ESS prediction performance in the training (*n* = 139) and holdout (*n* = 74) datasets. (*A*) and (*C*) are predicted change-in-ESS vs actual change-in-ESS plots, and (*B*) and (*D*) are Bland-Altman plots. Each point represents a single participant. The linear regression model was *Change-in-ESS = Baseline ESS + Baseline Event-Related Ventilatory Burden + Intercept* and was trained in the training dataset and evaluated in the holdout dataset. ESS = Epworth Sleepiness Scale score; *R²* = coefficient of determination between predicted change-in-ESS vs actual change-in-ESS; RMSE = root-mean-square error (interpreted as average change-in-ESS error per patient).

**Figure 6.**
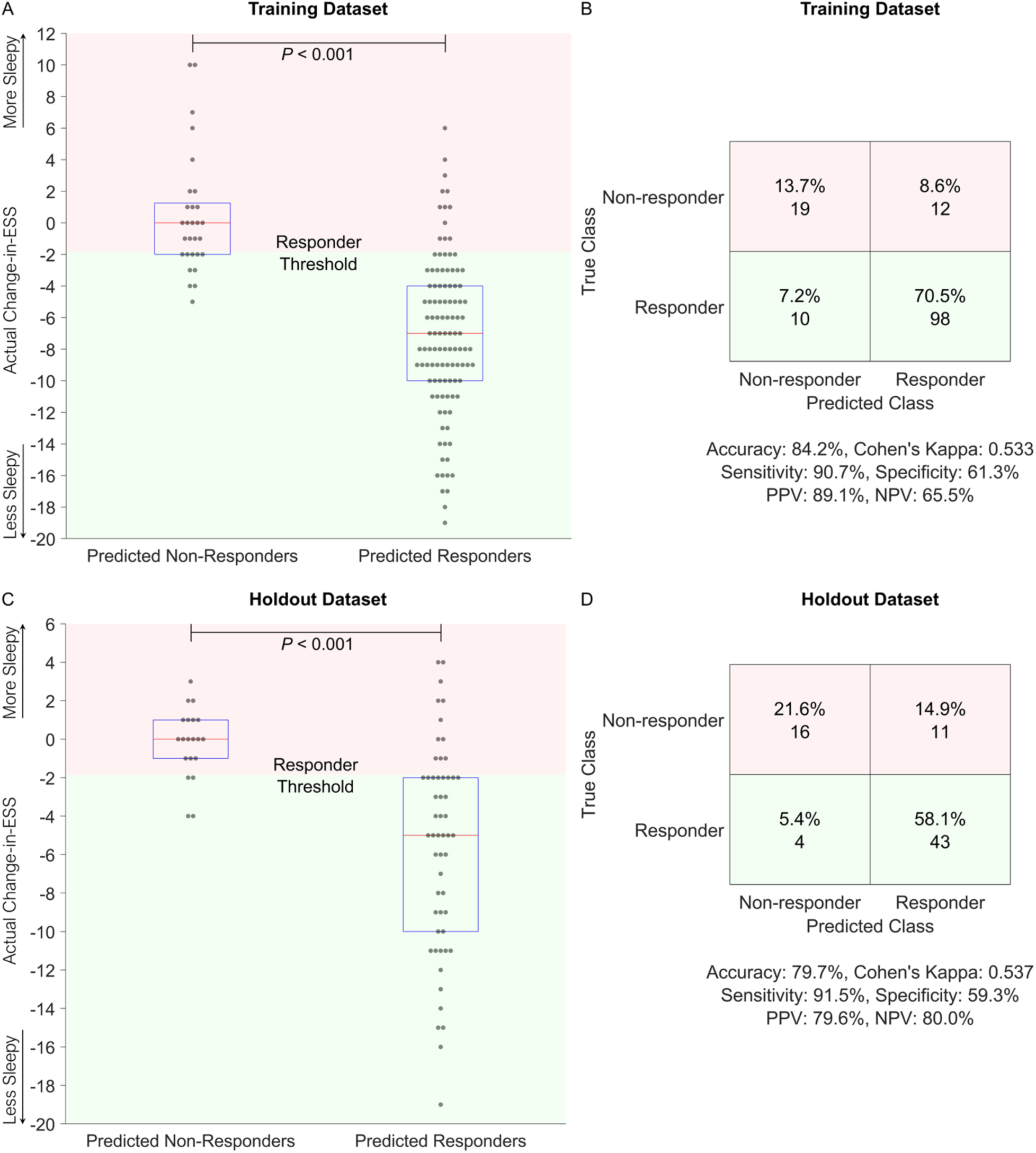
Boxplots and confusion matrices of results using the linear regression model as a classifier to predict responders in the training (*n* = 139) and holdout (*n* = 74) datasets. The change-in-ESS cut-point used to differentiate responders from non-responders was -2, which is the minimum clinically important difference of ESS in obstructive sleep apnea (21, 22). (*A*) and (*C*) are boxplots representing the 25^th^ percentile (lower horizontal line), median (red line), and 75^th^ percentile (upper horizontal line). Each participant is represented as a single point. The Mann-Whitney U-test was performed between predicted responder and non-responder groups. (*B*) and (*D*) are confusion matrices with summary performance metrics presented underneath. The model *Change-in-ESS = Baseline ESS + Baseline Event-Related Ventilatory Burden + Intercept* was developed in the training dataset and the optimal responder threshold was determined by the ROC convex hull method (50). False positives and false negatives were given a penalty weight of 1, and true positives and true negatives were given a penalty weight of 0. The model and optimal threshold were applied in the holdout dataset. ESS = Epworth Sleepiness Scale score; PPV = positive predictive value; NPV = negative predictive value.

**Table 3.**
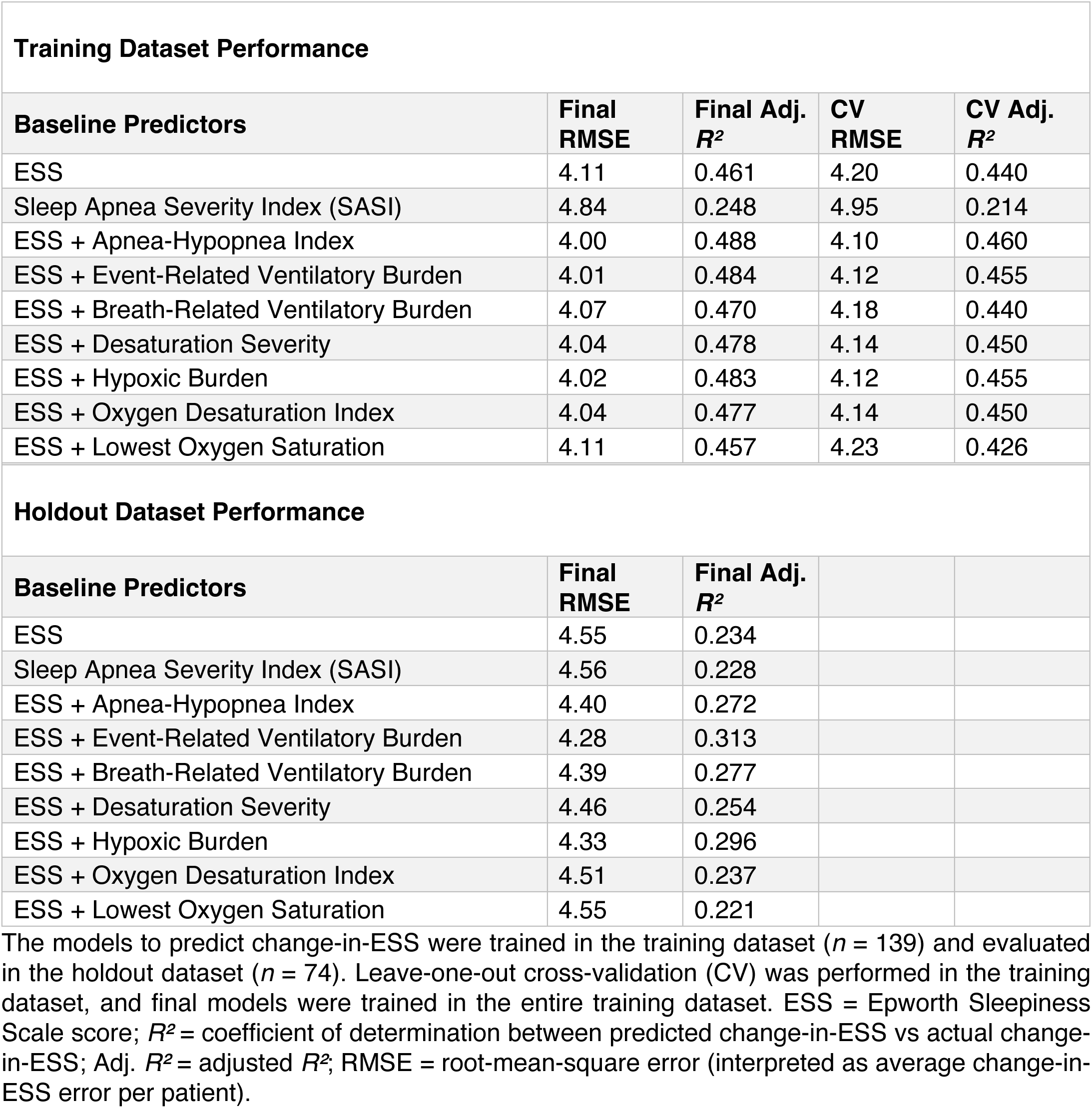
Performance of linear regression models that predict change in Epworth Sleepiness Scale score.

| Training Dataset Performance |  |  |  |  |
| --- | --- | --- | --- | --- |
| Baseline Predictors | Final RMSE | Final Adj. $R^2$ | CV RMSE | CV Adj. $R^2$ |
| ESS | 4.11 | 0.461 | 4.20 | 0.440 |
| Sleep Apnea Severity Index (SASI) | 4.84 | 0.248 | 4.95 | 0.214 |
| ESS + Apnea-Hypopnea Index | 4.00 | 0.488 | 4.10 | 0.460 |
| ESS + Event-Related Ventilatory Burden | 4.01 | 0.484 | 4.12 | 0.455 |
| ESS + Breath-Related Ventilatory Burden | 4.07 | 0.470 | 4.18 | 0.440 |
| ESS + Desaturation Severity | 4.04 | 0.478 | 4.14 | 0.450 |
| ESS + Hypoxic Burden | 4.02 | 0.483 | 4.12 | 0.455 |
| ESS + Oxygen Desaturation Index | 4.04 | 0.477 | 4.14 | 0.450 |
| ESS + Lowest Oxygen Saturation | 4.11 | 0.457 | 4.23 | 0.426 |
| Holdout Dataset Performance |  |  |  |  |
| Baseline Predictors | Final RMSE | Final Adj. $R^2$ | | |
| ESS | 4.55 | 0.234 |  |  |
| Sleep Apnea Severity Index (SASI) | 4.56 | 0.228 |  |  |
| ESS + Apnea-Hypopnea Index | 4.40 | 0.272 |  |  |
| ESS + Event-Related Ventilatory Burden | 4.28 | 0.313 |  |  |
| ESS + Breath-Related Ventilatory Burden | 4.39 | 0.277 |  |  |
| ESS + Desaturation Severity | 4.46 | 0.254 |  |  |
| ESS + Hypoxic Burden | 4.33 | 0.296 |  |  |
| ESS + Oxygen Desaturation Index | 4.51 | 0.237 |  |  |
| ESS + Lowest Oxygen Saturation | 4.55 | 0.221 |  |  |

The models to predict change-in-ESS were trained in the training dataset ( $n = 139$ ) and evaluated in the holdout dataset ( $n = 74$ ). Leave-one-out cross-validation (CV) was performed in the training dataset, and final models were trained in the entire training dataset. ESS = Epworth Sleepiness Scale score; $R^2$ = coefficient of determination between predicted change-in-ESS vs actual change-in-ESS; Adj. $R^2$ = adjusted $R^2$ ; RMSE = root-mean-square error (interpreted as average change-in-ESS error per patient).

A chart showing predicted change-in-ESS from baseline ventilatory burden and baseline ESS was created using the final linear regression model trained on all HomePAP, BestAIR, and ABC participants (Figure 7). In secondary exploratory analyses, %change-in-ESS was used as the outcome measure instead of change-in-ESS (Figures E8−E9 and Table E7).

**Figure 7.**
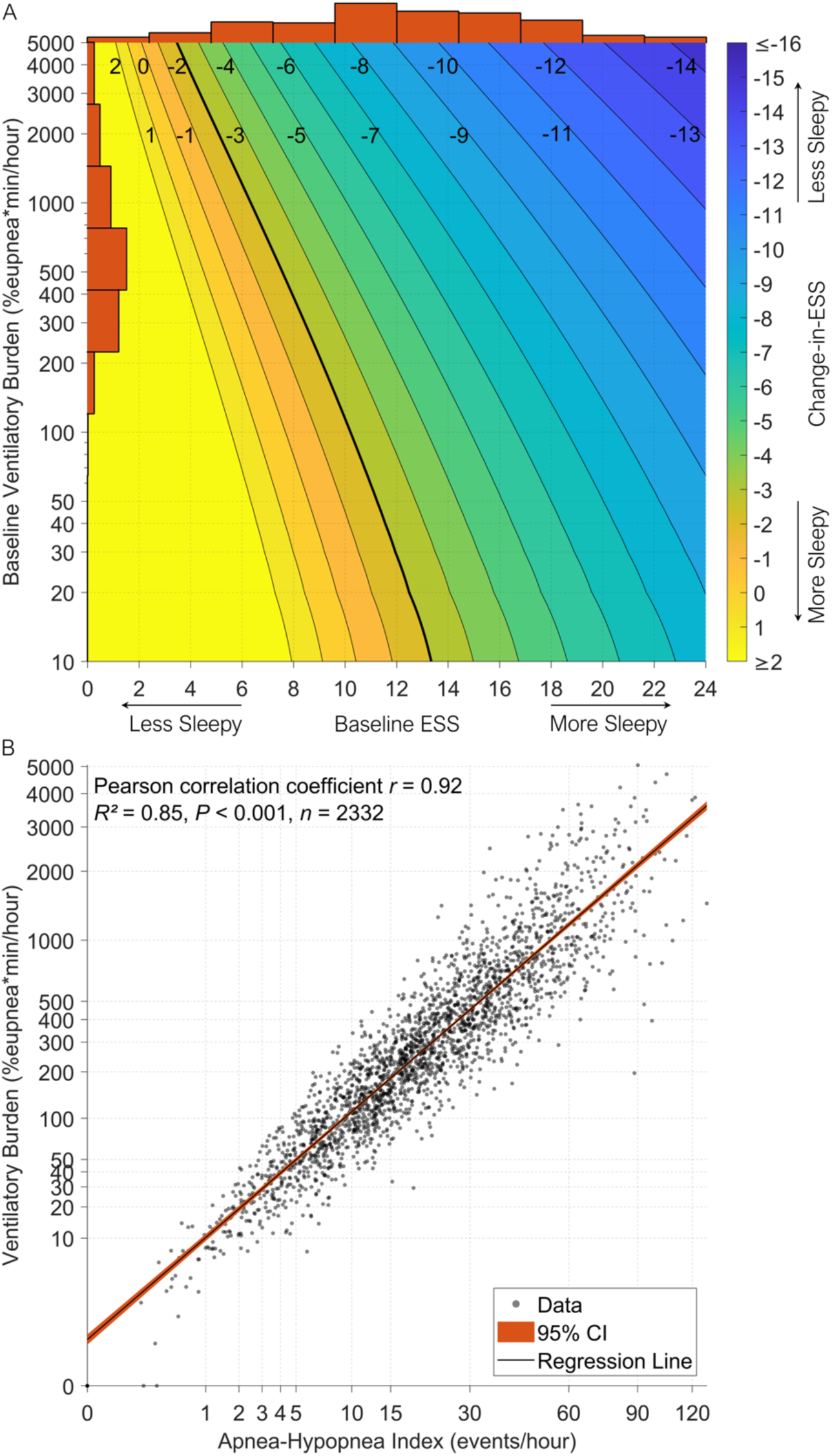
(*A*) Chart showing change-in-ESS predictions made by the final linear regression model trained on all HomePAP, BestAIR, and ABC participants (*n* = 213). Histograms along the top and left represent the distributions of baseline ESS and baseline event-related ventilatory burden in the 213 participants, respectively. The linear regression model was *Change-in-ESS = Baseline ESS + Baseline Event-Related Ventilatory Burden + Intercept*. The black contour lines with adjacent numbers represent change-in-ESS boundaries, and the thick black line represents change-in-ESS of -2 points, which is the minimum clinically important difference of ESS in obstructive sleep apnea (21, 22). As an example, a patient with baseline ESS of 8 points and baseline ventilatory burden of 400 %eupnea*min/hour is predicted to have a change-in-ESS of -2 points following CPAP treatment. (*B*) Scatter plot showing event-related ventilatory burden versus apnea-hypopnea index for 2332 participants from the combined cohort (black regression line with red shading representing 95% CI). The combined cohort consisted of HomePAP, BestAIR, ABC, STAGES, and MESA participants. ESS = Epworth Sleepiness Scale score.

## Discussion

The objective of this study was to determine whether characteristics from baseline polysomnography can predict change-in-ESS following CPAP treatment. First, associations between OSA severity measures and baseline ESS were evaluated to determine which variables were associated with sleepiness and might therefore be predictive of change-in-ESS. In 2332 participants, ESS was associated with ventilatory burden, hypoxic burden, AHI, and flow limitation severity, independent of potential confounders. Second, change-in-ESS prediction was evaluated using CPAP treatment studies (HomePAP, BestAIR, and ABC) in three steps: severity measures were compared (adjusted regression, *n*=64), a regression-based prediction model was developed in training participants (*n*=139), and then tested in holdout participants (*n*=74) to evaluate the model’s generalizability. Comparison analysis revealed that change-in-ESS was most strongly associated with baseline event-related ventilatory burden (i.e. cumulative lost ventilation per hour during sleep) and baseline ESS. These two variables were the most frequently chosen combination of predictors of change-in-ESS using cross-validated stepwise and LASSO selection of variables. The prediction model using baseline ventilatory burden and baseline ESS had higher correlation between predicted change-in-ESS and actual change-in-ESS than comparative models using baseline hypoxic burden and ESS; baseline AHI and ESS; baseline ESS only; and the Sleep Apnea Severity Index (SASI). The prediction model distinguished responders (≥2-point ESS improvement with CPAP) from non-responders with high sensitivity, low specificity, and slightly higher accuracy than the models trained using alternative metrics. Finally, the model was presented in the form of a chart designed to allow clinicians to predict change-in-ESS from baseline ventilatory burden and baseline ESS.

### Mechanistic insights

In this study, we demonstrated that ventilatory burden was associated with both baseline ESS and CPAP treatment-induced change-in-ESS. Ventilatory burden may contribute to hypersomnia in OSA via two pathways. First, lost ventilation during respiratory events may cause hypercapnia and acute respiratory acidosis leading to daytime sleepiness (27–29). Indeed, patients with OSA treated with CPAP show significant decreases in wake pCO_2_ and daytime sleepiness (30). Second, lost ventilation can lead to intermittent hypoxemia and hypoxia, leading to oxidative injury, changes in neuronal connectedness, and increased inflammation in wake-promoting regions of the brain and brainstem (31, 32). We also observed that hypoxic burden was associated with baseline ESS and change-in-ESS, which supports this notion. However, the effect size of ventilatory burden was slightly larger than that of hypoxic burden, possibly because ventilatory burden captures both hypoxemia and hypercapnia aspects of OSA.

Several studies have examined the relationship between baseline polysomnographic characteristics and change-in-ESS following CPAP treatment. Bhat *et al.* reported significant correlations between change-in-ESS and baseline AHI and T90 in severe OSA (8). However, only patients who were adherent (≥4 hours CPAP use/night for 60% of nights) and adequately treated (residual AHI≤5 events/hour) were included in their study. The results of our study – which included all individuals regardless of adherence and residual AHI – are in concordance, i.e. higher baseline OSA severity (ventilatory burden) was associated with greater ESS improvement. In contrast, Kingshott *et al.* found no significant correlation between change-in-ESS and baseline AHI, arousal index, ODI, and minimum oxygen saturation in severe OSA (9). They did, however, find that CPAP use (hours/night) was associated with change-in-ESS. Similarly, Otsuka *et al.* showed that baseline AHI was not a predictor of change-in-ESS and that CPAP use was (10). Other studies have demonstrated a dose-response relationship between CPAP usage and improvement in self-reported sleepiness (33, 34). This contrasts with our findings that CPAP use was not significantly associated with the change-in-ESS (Figure E3). Likewise, prior analysis of the HomePAP cohort (35) indicated that CPAP was associated with change-in-ESS, but not after adjusting for baseline AHI, and that higher baseline AHI was associated with ESS normalization. Furthermore, Bonsignore *et al.* showed that change-in-ESS was weakly correlated with CPAP use (*R²*=0.023), and baseline AHI and oxygen saturation parameters were meaningfully different between patients with and without hypersomnia at baseline and follow-up (7). Thus, there is now considerable evidence that baseline disease severity, as well as adherence, influences the improvement in sleepiness in patients treated with CPAP.

We observed that flow limitation was a significant predictor of ESS in the cross-sectional analysis, which is consistent with earlier studies (18, 20, 36). However, flow limitation severity and frequency were not significant predictors of change-in-ESS in the HomePAP home cohort. The explanation for this may lie with the severity of OSA; flow limitation appears to be most strongly associated with sleepiness in patients without severe OSA (20), and the HomePAP trial participants had relatively severe OSA. Flow limitation may therefore fail to adequately summarize the severity of breathing disturbance in more severe OSA. Nonetheless, in the absence of more severe OSA, flow limitation appears to contribute to OSA-related symptoms: studies have demonstrated that sustained inspiratory flow limitation from suboptimal CPAP pressure was associated with decreased vigilance (37), and that elimination of residual flow limitation with CPAP improves vigilance (38).

### Clinical implications

The American Academy of Sleep Medicine (AASM) International Classification of Sleep Disorders states that to diagnose adult OSA, a patient must have either AHI≥15 events/hour or AHI≥5 events/hour and one or more symptoms, e.g. sleepiness (39). AASM guidelines for the treatment of adult OSA recommend that clinicians should use CPAP to treat OSA in adults with hypersomnia (40). The ESS is the most common quantification of sleepiness in OSA, with scores >10 indicating hypersomnia (3). From this guidance, clinicians should prescribe CPAP to patients with AHI≥5 and ESS>10. However, we found that change-in-ESS following CPAP treatment is modulated independently by baseline ventilatory burden and baseline ESS. From our chart, there may be patients with baseline ESS≤10 points who may have a clinically meaningful improvement in sleepiness (≥2-point ESS improvement (21, 22)) with CPAP treatment but would not be prescribed CPAP based on clinical guidelines (Figure 7). For example, a patient with baseline ESS=8 points, ventilatory burden=400 %eupnea*min/hour, and AHI=10 events/hour is expected to have a 2-point ESS improvement. Conversely, there may be patients with AHI≥5 and ESS>10 who are prescribed CPAP but do not have a meaningful improvement in sleepiness due to low ventilatory burden. For example, a patient with baseline ESS=11 points, ventilatory burden=30 %eupnea*min/hour, and AHI=10 events/hour is expected to have a 1-point ESS improvement. Therefore, our modeling approach has promise as a means to prescribe CPAP more judiciously than current clinical guidelines.

We have addressed several considerations that should be made to develop clinically useful prediction models (41). Our model addresses a clear clinical decision point: whether to prescribe CPAP for OSA. The model outputs expected change-in-ESS with a confidence interval and our easy-to-use chart helps clinicians make this decision. The model’s input parameters are baseline ESS and baseline ventilatory burden, which can be calculated from routine polysomnograms using publicly available code (12), making it feasible to integrate into clinical data collection and reporting systems. The AHI could be used in place of ventilatory burden (with a modest loss of performance) when ventilatory burden is unfeasible to calculate. In addition to predicting continuous change-in-ESS, our model can be used to identify likely responders/non-responders with high sensitivity (91.5%) and low specificity (59.3%), which reflects the current clinical practice of prescribing CPAP (35). We showed that 79.6% of predicted responders were actual responders (Figure 6D); if all predicted responders were treated with CPAP there would have been a 44.2% reduction in the rate of non-response compared to treating all patients. If predicted change-in-ESS was available clinically, clinicians and patients could potentially make more informed decisions about the likelihood by which the intervention would improve sleepiness.

Our study implicates ventilatory burden as a candidate measure for OSA severity in place of the AHI. Ventilatory burden is the product of average respiratory event depth, average event duration, and event frequency (Figure 1). Thus, for interpreting the effects of OSA on OSA-related symptoms, respiratory event depth and duration may also be important, beyond event frequency. Ventilatory burden also appeared to perform better than the leading comparator for predicting treatment response, i.e. the “SASI”, which has been associated with both change in PSQI and change in Sleep Apnea Quality of Life Index (11). The SASI is calculated using baseline ESS, AHI, BMI, lowest oxygen saturation, and redundant pharyngeal mucosa (which was not measured and assumed absent for all participants in our study). We found that the SASI was not sufficiently correlated with change-in-ESS (Pearson’s *r*=-0.52, Figure E2) to provide a useful cross-validated prediction of change-in-ESS and responders/non-responders. Thus, overall ventilatory burden may be a better disease severity measure of OSA than both the AHI and SASI, passing necessary tests: 1) a cross-sectional association with sleepiness, 2) an association with the treatment-related improvement in sleepiness, and 3) demonstrated ability to predict improvement in sleepiness.

### Methodological considerations

This study has several limitations. First, we consider that ventilatory burden may have appeared superior to hypoxemia measures potentially due to heterogeneity in pulse oximetry technology across studies. Thus, the utility of hypoxemia measures as a prediction tool may be somewhat compromised by differences across sites/devices; standardization of technology may improve their predictive ability. Second, the ESS is a subjective measurement of sleepiness. However, in support of the ESS, it has been found to have greater discriminating power than the maintenance of wakefulness test (MWT) and multiple sleep latency test (MSLT) for narcolepsy (42). Additionally, Sun *et al.* observed that functional brain changes after CPAP treatment could only be found by grouping participants based on ESS but not MSLT (43). Furthermore, the ESS is used ubiquitously in sleep medicine, has high internal consistency (44), and the MWT and MSLT were not administered in HomePAP, BestAIR, and ABC studies. Third, baseline ventilatory burden and baseline ESS could not fully explain all variance in change-in-ESS with CPAP treatment. CPAP resolves airway obstruction; however, a patient might be sleepy for reasons other than OSA, e.g. insufficient sleep duration, comorbidities such as obesity and other sleep or circadian disorders, and medications (32, 45). We found that total sleep time and baseline BMI were not significant predictors of change-in-ESS; medication use data was not available in the CPAP treatment studies. Furthermore, heterogeneity in OSA-related neuronal damage via long-term exposure to intermittent hypoxia (46–48) may explain the inability to fully predict change-in-ESS. Fourth, the median follow-up duration in HomePAP was three months; larger ESS improvements may have been observed if the follow-up duration was longer. Indeed, Bonsignore *et al.* reported that the prevalence of hypersomnia decreased as the first follow-up time point after CPAP treatment initialization was extended beyond 3 months (7). Furthermore, change-in-ESS prediction may have been more accurate if the follow-up period was consistent across studies. Fifth, we did not quantify sleep fragmentation and arousal parameters because electroencephalography was not recorded in HomePAP and BestAIR home polysomnograms. However, previous work in a cross-sectional cohort shows that arousal severity was less strongly associated with ESS compared to flow limitation, hypoxemia, and AHI (18). Sixth, no control group was used in this analysis; patient expectation of CPAP benefit can influence change-in-ESS, however, this placebo effect is expected to be approximately -1.2 points (49). Finally, follow-up polysomnograms were either not collected or unusable in the CPAP treatment studies which prevented us from quantifying changes in polysomnography-derived metrics and their relationships with change-in-ESS.

## Conclusions

Our work provides novel insight into the polysomnographic measures that are most predictive of the CPAP-related improvement in sleepiness in patients with OSA. Baseline ventilatory burden and baseline ESS were independently associated with change-in-ESS and could be used together to inform clinicians whether CPAP treatment will likely improve a patient’s sleepiness.

## Data Availability

This study used data from five published studies that are publicly available from the National Sleep Research Resource website:
Apnea, Bariatric surgery, and CPAP study (ABC): https://sleepdata.org/datasets/abc
Best Apnea Interventions in Research (BestAIR): https://sleepdata.org/datasets/bestair
Home Positive Airway Pressure (HomePAP): https://sleepdata.org/datasets/homepap
Stanford Technology Analytics and Genomics in Sleep (STAGES): https://sleepdata.org/datasets/stages
Multi-Ethnic Study of Atherosclerosis (MESA): https://sleepdata.org/datasets/mesa
Requests for code and processed data should be directed to the corresponding author.

## Data sharing statement

This study used data from five published studies that are publicly available from the National Sleep Research Resource website:

- Apnea, Bariatric surgery, and CPAP study (ABC): https://sleepdata.org/datasets/abc
- Best Apnea Interventions in Research (BestAIR): https://sleepdata.org/datasets/bestair
- Home Positive Airway Pressure (HomePAP): https://sleepdata.org/datasets/homepap
- Stanford Technology Analytics and Genomics in Sleep (STAGES): https://sleepdata.org/datasets/stages
- Multi-Ethnic Study of Atherosclerosis (MESA): https://sleepdata.org/datasets/mesa

Requests for code and processed data should be directed to the corresponding author.

## Clinical trial statement

This was a secondary analysis of retrospective data. Some of the data utilized originated from clinical trials registered with www.clinicaltrials.gov (NCT00642486, NCT01261390, and NCT01187771).

### Acknowledgements

The authors thank the staff of the National Sleep Research Resource and the researchers and participants of the studies listed below. The National Sleep Research Resource was supported by the U.S. National Institutes of Health, National Heart Lung and Blood Institute (R24 HL114473, 75N92019R002).

The Apnea, Bariatric surgery, and CPAP study (ABC Study) was supported by National Institutes of Health grants R01HL106410 and K24HL127307. Philips Respironics donated the CPAP machines and supplies used in the perioperative period for patients undergoing bariatric surgery.

The Best Apnea Interventions in Research (BestAIR) was supported by the National Heart, Lung, and Blood Institute (1U34HL105277) and a grant from ResMed Foundation. Equipment was donated by ResMed and Philips Respironics.

The Home Positive Airway Pressure study (HomePAP) was supported by the American Sleep Medicine Foundation 38-PM-07 Grant: Portable Monitoring for the Diagnosis and Management of OSA.

The Multi-Ethnic Study of Atherosclerosis (MESA) Sleep Ancillary study was funded by NIH-NHLBI Association of Sleep Disorders with Cardiovascular Health Across Ethnic Groups (RO1 HL098433). MESA was supported by NHLBI funded contracts HHSN268201500003I, N01-HC-95159, N01-HC-95160, N01-HC-95161, N01-HC-95162, N01-HC-95163, N01-HC-95164, N01-HC-95165, N01-HC-95166, N01-HC-95167, N01-HC-95168 and N01-HC-95169 from the National Heart, Lung, and Blood Institute, and by cooperative agreements UL1-TR-000040, UL1-TR-001079, and UL1-TR-001420 funded by NCATS.

This research has been conducted using the STAGES (Stanford Technology, Analytics and Genomics in Sleep) Resource funded by the Klarman Family Foundation. The investigators of the STAGES study contributed to the design and implementation of the STAGES cohort and/or provided data and/or collected biospecimens, but did not participate in the analysis or writing of this report. The full list of STAGES investigators can be found at the project website (https://sleepdata.org/datasets/stages).

