## Supplementary Information for "Ventilatory Burden Predicts Change in Sleepiness Following Positive Airway Pressure in Sleep Apnea"

### Supplementary Methods

<u>STAGES polysomnographic recordings</u>

In the STAGES cohort, polysomnograms were scored according to the 2007 American Academy of Sleep Medicine (AASM) guidelines (1). Respiratory events were not filtered because manually scored arousal data was not available; if respiratory events had been filtered, this may have led to discarding hypopneas with associated arousals.

<u>MESA polysomnographic recordings</u>

In the MESA cohort, hypopneas were scored regardless of desaturation using AASM 30% recommended and 50% alternative criteria (2).

<u>HomePAP polysomnographic recordings</u>

HomePAP studies were scored according to AASM criteria of 30% reduction in nasal airflow signal associated with at least a 4% desaturation (3). HomePAP participants were randomized to either home or in-lab diagnostic polysomnography. Embletta polysomnography recording equipment was used for home studies. Participants with a home study AHI<15 events/hour also had an in-lab polysomnography to determine their eligibility to participate in the study. In the analyses herein, the AHI for patients who had both home and in-lab polysomnography was calculated from their in-lab study.

In HomePAP home polysomnograms, sleep staging was not performed and total sleep time (TST) could not be computed because electroencephalography was not recorded. However, total recording time was estimated from channels other than electroencephalography using a validated method (4). Of the initial 373 HomePAP participants, 63 participants had both home and in-lab polysomnography as well as total analyzable flow time ≥2 hours. The median ratio between home polysomnography total recording time and in-lab polysomnography TST was 1.298 for these participants. The 5$^{th}$ and 95$^{th}$ percentiles of the ratio values were 1.015 and 2.010, respectively. This scaling factor was used to adjust home polysomnography total recording time to be within the range of laboratory polysomnography TST. This TST surrogate was used in subsequent calculations of the AHI, desaturation severity, event-related ventilatory burden, hypoxic burden, and oxygen desaturation index (ODI).

<u>BestAIR polysomnographic recordings</u>

BestAIR sleep studies were either performed at home ($n$ = 57) or in-lab ($n$ = 5). Embletta polysomnography recording equipment was used for home studies. When arousal data was available, polysomnograms were scored according to AASM alternative hypopnea criteria; otherwise, they were scored using the AASM recommended criteria when electroencephalographic data was not recorded (5). Wake and sleep were manually scored in the Type 3 home studies based on channels other than electroencephalography. TST was calculated from the manual scoring of sleep. Sleep stages and arousals were manually scored in the Type 1 in-lab studies. Some of the in-lab studies were split-night CPAP titration; throughout the titration period, sleep stages were automatically set to wake, and the nasal airflow signal was discarded. Breath-related ventilatory burden during sleep including arousals was computed because the home studies had sleep/wake scoring but no arousal scoring.

ABC polysomnographic recordings

All analyzed ABC polysomnograms were Type 1 and scored according to AASM criteria (6).

Respiratory and desaturation event filtering

All polysomnograms from all cohorts were scored according to the AASM guidelines; however, different criteria were used between cohorts. Inconsistent criteria to define apneas, hypopneas, and oxygen desaturations is an issue when defining and categorizing obstructive sleep apnea (OSA) severity (7). Therefore, in this study, scored respiratory and desaturation events were filtered to make scoring consistent across cohorts. Only respiratory events that began in an epoch of sleep were included. All apneas were included and only hypopneas with ≥3% desaturation or arousal were kept (AASM recommended criteria), which is consistent with current AASM guidelines (8). Only desaturations ≥3% from baseline to nadir were kept.

CPAP adherence

CPAP adherence data (nightly CPAP use) was only available in HomePAP and not available in BestAIR and ABC studies.

Epworth Sleepiness Scale score (ESS) and other follow-up measurements

Change-in-ESS was calculated from untransformed follow-up ESS and untransformed baseline ESS (i.e. follow-up ESS – baseline ESS) and then transformed using the Box-Cox transformation (see "Statistical analyses" section below). In HomePAP, follow-up ESS, BMI, and nightly CPAP use was recorded at 1 month and 3 months after treatment initiation. Data from the 3-month follow-up was preferentially used. 15 participants did not have this data, so their 1-month follow-up data was used instead. In BestAIR, follow-up ESS and BMI was recorded at 6 months and 12 months after treatment initiation. Data from the 6-month follow-up was preferentially used to be closer to the 3-month follow-up from HomePAP studies. Two participants did not have this data, so their 12-month follow-up data was used instead. In ABC, follow-up ESS and BMI was recorded at 9 months and 18 months after treatment initiation. Data from the 9-month follow-up was preferentially used to be closer to the 3-month follow-up from HomePAP studies. All participants had 9-month follow-up ESS data.

Computing ventilatory burden

"Event-related ventilatory burden" is cumulative lost ventilation per hour during sleep and was computed using the uncalibrated nasal airflow signal and scored respiratory events, as described (9, 10). Filtered respiratory events using AASM recommended criteria were used. Periods of useable nasal airflow signal were manually marked, and all unusable periods were discarded. Breaths were automatically detected, and tidal volume was estimated by integrating the nasal airflow signal. Eupneic ventilation was considered the mean tidal volume per 7-minute sliding window (2-minute steps). Minute ventilation on a breath-by-breath basis was calculated based on eupneic ventilation. The depth of lost ventilation was calculated for each respiratory event. Event-related ventilatory burden was calculated from: average respiratory event depth × average respiratory event duration × respiratory event frequency. The units of event-related ventilatory burden are %eupnea*min/hour. Two minor modifications were made to the published event-related ventilatory burden algorithm (9): first, the average respiratory event depth under 90% eupnea was used instead of the average depth under 90% of the ensemble averaged respiratory event ventilation signal; second, average event duration was calculated by averaging the duration of ventilation under 90% eupnea for each event instead of averaging manually scored event durations.

"Breath-related ventilatory burden" is the percentage of breaths during sleep under 50% eupnea, modified from Parekh *et al.* (11). Like flow limitation metrics, all breaths during sleep excluding arousal breaths were used to compute breath-related ventilatory burden. In STAGES, all breaths during sleep including arousals were used because arousals were not scored. In HomePAP home studies, all breaths during the study were used because sleep stages and arousals could not be scored.

Both ventilatory burden measures were quantified in all cohorts – even those with high-pass filtered and low signal-to-noise ratio (SNR) nasal airflow – because ventilation measurements are tolerant to noisy nasal airflow signal (10).

<u>Computing flow limitation</u>

We used two validated algorithms to objectively assess the degree of flow limitation on a breath-by-breath basis using the nasal airflow signal. Flow limitation frequency is defined as the percentage of breaths during sleep classified as "certain flow-limited" (12). This classification was based on multiple flow shape characteristics (e.g., fluttering and scooping) that describe non-rounded inspiratory/expiratory flow (12). We also assessed flow limitation severity, which quantifies the mismatch between measured ventilation and intended ventilation (ventilatory drive) (13). Specifically, flow limitation severity = (1 – (ventilation ÷ drive)) × 100. A score of 50% indicates ventilation was half of the intended level, and a score of 0% indicates a perfectly patent airway. The median value of flow limitation severity provided a single summary metric (13). All breaths during sleep including those that occurred within scored respiratory events were included in analyses. Breaths within obstructive apneas – scored apneas plus any unscored periods of ventilation below 10% of the local mean value (7-minute sliding window) – were imputed at the median respiratory rate and attributed a "certain flow-limited" label and a flow limitation severity of 99.9%. Breaths were excluded if they were within scored arousals or longer than Q3 + 1.5 × IQR (where Q3 = quartile 3 and IQR = interquartile range of a participant's breath durations).

Flow limitation frequency and severity were not computed in ABC and HomePAP in-lab studies which had high-pass filtered nasal airflow data, or BestAIR polysomnograms which had low nasal airflow signal-to-noise ratio.

Arousal scoring was not available in the STAGES cohort polysomnograms, so flow limitation severity during all sleep epochs including arousal breaths was quantified instead. In STAGES studies, a published correction factor was used to correct flow limitation severity based on estimated signal-to-noise ratio (10, 14). Flow limitation frequency was not quantified in STAGES because no correction factor has been developed for this algorithm yet.

To quantify flow limitation severity in MESA, a model trained on 10Hz flow-shape features was used (15). For flow limitation frequency, the original flow limitation certainty model was used because using 10Hz flow-shape features was not inferior to using 25Hz flow-shape features (15).

Sleep stage and arousal scoring was not available in HomePAP home studies because electroencephalography was not recorded. Therefore, flow limitation for all breaths during the polysomnograms was quantified. In the MESA cohort, there was a very strong correlation between flow limitation frequency excluding arousal breaths and flow limitation frequency for all breaths (Pearson's $r = 0.94$). Similarly, the correlation between flow limitation severity excluding arousal breaths and flow limitation severity for all breaths was high ($r = 0.81$).

Computing hypoxemia severity

The oxygen saturation signal and filtered respiratory events using AASM recommended criteria were used to automatically find resulting desaturations and compute hypoxic burden (16). The original method was modified to improve desaturation baseline detection by using a search window for the desaturation baseline instead of setting each desaturation baseline to be the maximum SpO2 value 100 seconds prior to the end of each respiratory event. The left search window was set to be the ensemble averaged circulatory delay minus half the respiratory event duration. The right search window was set to be the ensemble averaged circulatory delay plus the ensemble averaged time from start of desaturation to nadir. The following three modifications were made when finding each desaturation: 1) prevent setting the desaturation baseline before the previous desaturation's nadir SpO2 value, achieved by capping the baseline search window left-hand side to be at the end of the previous respiratory event if the baseline search window left-hand side occurred before the end of the previous respiratory event; 2) prevent setting the baseline to be the baseline of the next desaturation, achieved by capping the baseline search window right-hand side to be at the start of the next respiratory event if the baseline search window right-hand side occurred after the start of the next respiratory event; and 3) prevent setting the baseline to be after the nadir, achieved by capping the baseline search window right-hand side to be at the desaturation nadir if the baseline search window right-hand side occurred after the desaturation nadir. The start of each automatically identified desaturation was defined as the maximum SpO2 value within the baseline search window. The end of each desaturation was defined as the maximum point from the desaturation nadir to the right-hand side of the full search window. For the purposes of calculating desaturation area, the baseline was defined as the maximum SpO2 value within the identified desaturation. The area of each desaturation was calculated by integrating the SpO2 signal from this baseline within the identified desaturation start and end points. Hypoxic burden was calculated by taking the average area of all desaturations and multiplying by the AHI.

The oxygen saturation signal and filtered desaturations using ≥3% criteria were used to compute desaturation severity (17) and oxygen desaturation index (ODI). Desaturation severity and ODI were not quantified in the STAGES cohort because desaturation scoring was not available in polysomnograms recorded at the Stanford Sleep Medicine Center (Redwood City, United States).

Computing the Sleep Apnea Severity Index (SASI)

The Sleep Apnea Severity Index (SASI) was calculated as described (18) using baseline ESS, AHI, BMI, and lowest oxygen saturation. The three possible categories are mild (I), moderate (II), and severe (III). The SASI is also calculated using redundant pharyngeal mucosa and modified the SASI uses tonsil size instead (19). Neither of these measurements were available in any of the cohorts analyzed in this study, so redundant pharyngeal mucosa was assumed absent for all participants when computing the SASI. The SASI was not included as a predictor variable in multivariable statistical inference analyses because the SASI itself is a multivariable model.

Electroencephalography-based metrics

Electroencephalography-based metrics were not computed because electroencephalography was not recorded in HomePAP and BestAIR home (Type 3) polysomnograms.

Statistical analyses

One STAGES participant was missing ESS data; one HomePAP in-lab participant and three MESA participants were missing BMI data; and hypoxic burden could not be quantified in four participants. One HomePAP participant was missing data on nightly CPAP usage. Depression data was missing in three HomePAP home participants, three HomePAP in-lab participants, and four BestAIR participants.

Data processing and statistical analyses were performed in MATLAB (version R2024a, Mathworks, Natick, MA, USA) and R (version 4.2.2, R Foundation, Vienna, Austria). All continuous variables were transformed to be normally distributed using the two-parameter Box-Cox (20) transformation. Transformed values were used in all subsequent analyses. Back-transformation was used to show values in original units of measurement. Box-Cox transformation parameters were derived in all participants from all studies and then applied to all studies. The two-parameter Box-Cox transformation is $Y' = (Y+\lambda_2)^{\lambda_1}-1) \div \lambda_1$ or $Y' = \log_e(Y+\lambda_2)$ if $\lambda_1 = 0$. Optimal values of $\lambda_1$ and $\lambda_2$ were estimated using the boxcoxfit function from the geoR package (version 1.9-2) of R (21, 22). After transformation, Y' was standardized between 0 and 1 by subtracting the minimum value and dividing by the range. Values of $\lambda_1$, $\lambda_2$, minimum value, range, and sample size are presented in Table E1 for each variable and may be used to transform variables in other studies, provided they are in the ranges shown in Figure E2. For parameters such as total sleep time, $n$ = 2332 participants were used to compute transformation parameters. For parameters such as change-in-ESS, $n$ = 213 participants were used because this variable could only be computed using HomePAP, BestAIR, and ABC studies.

Change-in-ESS prediction model training and evaluation

Change-in-ESS prediction was evaluated using HomePAP, BestAIR, and ABC studies. The HomePAP home cohort was used for variable selection because the polysomnograms from these studies had unfiltered and high SNR nasal airflow which is necessary to compute flow limitation metrics. Then, all HomePAP home participants and 50% of HomePAP in-lab, BestAIR, and ABC participants (allocated using every second participant identification number, total $n$ = 139) were used to train a linear regression model to predict change-in-ESS based on baseline ventilatory burden and baseline ESS. The training dataset consisted of a mix of sleepy and non-sleepy participants and a mix of home and in-lab studies. This model was then tested in a holdout dataset consisting of the remaining 50% of HomePAP in-lab, BestAIR, and ABC participants (total $n$ = 74).

Stepwise linear regression was used to determine the best predictors of change-in-ESS. The “stepwiselm” MATLAB function was used and parameters were set to: criterion = “sse”; maximum steps = 1000; initial terms = constant term only; upper bound on terms = linear; threshold for adding a term = $P < 0.05$; and threshold for removing a term = $P > 0.1$. Leave-one-out cross-validation was performed, and final models were trained in all HomePAP home participants.

We also performed LASSO selection of variables and compared performance and selected variables against stepwise linear regression. LASSO is a regression method that incorporates variable selection and regularization (i.e. sensible simplification to avoid overfitting). The “lasso” MATLAB function was used with alpha set to 1. The optimal lambda value was found by running the “lasso” function with leave-one-out cross-validation (by setting the number of cross-validation folds to the sample size, i.e. 64) and using the lambda value with minimum cross-validation error. Then, leave-one-out cross-validation was performed using the optimal lambda value for the “Lambda” parameter and “resubstitution” for the “CV” parameter.

Linear regression change-in-ESS prediction performance was evaluated in the training and holdout datasets. In exploratory analyses, we compared the predictive performance of alternative linear regression models using other sets of predictor variables. A chart showing change-in-ESS predictions was generated using the final linear regression model trained on all HomePAP, BestAIR, and ABC participants.

The linear regression model *Change-in-ESS = Baseline ESS + Baseline Event-Related Ventilatory Burden + Intercept* trained on the training dataset was used as a classifier to predict responders in both the training and holdout datasets. The change-in-ESS cut-point used to differentiate responders from non-responders was -2 based on the minimum clinically important difference of ESS in OSA (23, 24). A receiver operating characteristic (ROC) curve was generated and the optimal responder probability threshold was determined by the ROC convex hull method (25). False positives and false negatives were given an equal penalty weight of 1, and true positives and true negatives were given a penalty weight of 0. The model and optimal threshold were applied in the holdout dataset. In sensitivity analyses, false positives were given a penalty weight of 0.5 and false negatives were given a penalty weight of 1 to reflect the current clinical practice of overprescribing rather than under-prescribing CPAP (26).

### Supplementary Figure E1 (Participant Exclusion Criteria)

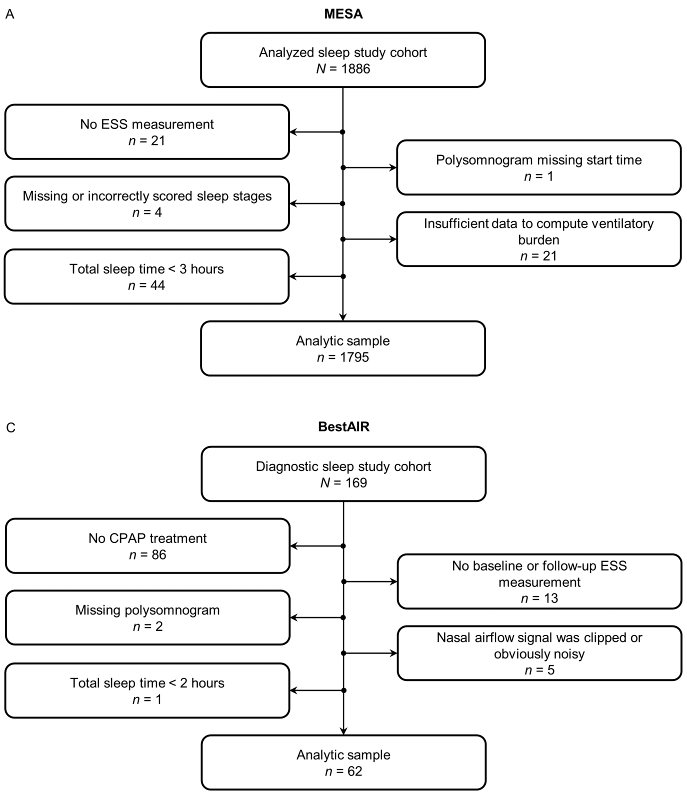

**Supplementary Figure E1.** Flow diagram of participant exclusion criteria applied to the diagnostic sleep studies performed in the (*A*) MESA cohort, (*B*) HomePAP cohort, (*C*) BestAIR cohort, and (*D*) ABC cohort. The derivation of the MESA "analyzed sleep study cohort" (*n* = 1886) from the MESA "sleep exam cohort" (*n* = 2237) was reported by Mann *et al.* (15). Participant exclusion criteria applied to the STAGES cohort has been reported (14). ESS = Epworth Sleepiness Scale score; CPAP = continuous positive airway pressure; *n* = sample size.

**Supplementary Table E1 (Box-Cox Parameters)**

**Supplementary Table E1.** Box-Cox transformation values of continuous variables in the combined cohort consisting of all participants from HomePAP, BestAIR, ABC, STAGES, and MESA cohorts.

| | $\lambda_1$ | $\lambda_2$ | Minimum value | Range | Sample size (*n*) |
|---|---|---|---|---|---|
| **Age (years)** | 2.0568 | 0 | 110.2221 | 5.3295 × 10^3 | 2332 |
| **Baseline Body Mass Index (kg/m²)** | -0.9892 | 4.8074 | 0.9614 | 0.0357 | 2328 |
| **Follow-up Body Mass Index (kg/m²)** | -0.4596 | 0 | 1.6329 | 0.2330 | 194 |
| **Total Sleep Time (hours)** | 5.9171 | 65.8969 | 1.1690 × 10^10 | 1.2195 × 10^10 | 2332 |
| **Apnea-Hypopnea Index (events/hour)** | 0.3199 | 0.0013 | -2.7540 | 14.4229 | 2332 |
| **Nasal Airflow Signal-to-Noise Ratio (dB)** | -4.5768 | 228.1822 | 0.2185 | 2.6799 × 10^-12 | 2330 |
| **Flow Limitation Frequency (%)** | 0.1832 | 9.2449 × 10^-4 | -3.9402 | 10.9917 | 1859 |
| **Flow Limitation Severity (%)** | -4.0396 | 173.1051 | 0.2476 | 1.6942 × 10^-10 | 2183 |
| **Event-Related Ventilatory Burden (%eupnea*min/hour)** | 0.1718 | 0.0506 | -2.3351 | 21.7049 | 2332 |
| **Breath-Related Ventilatory Burden (%)** | 0.0458 | 0.0403 | -2.9857 | 8.0389 | 2332 |
| **Desaturation Severity (%)** | 0.0910 | 0.0289 | -3.0297 | 6.9444 | 2008 |
| **Hypoxic Burden (%min/hour)** | 0.2092 | 0.0118 | -2.8917 | 19.1060 | 2328 |
| **Oxygen Desaturation Index (events/hour)** | 0.2952 | 0.0020 | -2.8483 | 15.6023 | 2332 |
| **Lowest Oxygen Saturation (%)** | 3.2151 | 0 | 4.4019 × 10^4 | 6.9063 × 10^5 | 2332 |
| **Nightly CPAP Use (hours)** | 0.5605 | 8.8000 × 10^-5 | -1.7747 | 6.0272 | 128 |
| **Baseline ESS (points)** | 0.4857 | 2.4000 × 10^-4 | -2.0231 | 9.6020 | 2331 |
| **Follow-up ESS (points)** | 0.3346 | 0 | 0 | 5.2890 | 213 |
| **Change-in-ESS (points)** | 0.7669 | 19.0000 | -1.3039 | 17.2499 | 213 |
| **Change-in-ESS (%)** | 0.0741 | 107.4830 | 2.7758 | 4.9178 | 213 |

The two-parameter Box-Cox transformation is $Y' = (Y+\lambda_2)^{\lambda_1}-1) \div \lambda_1$ or $Y' = \log_e(Y+\lambda_2)$ if $\lambda_1 = 0$. After transformation, Y' was standardized between 0 and 1 by subtracting the minimum value and dividing by the range. Not all variables were measured in all cohorts, e.g. follow-up ESS was not measured in STAGES and MESA cohorts due to their cross-sectional nature which prevented calculation of change-in-ESS. In cross-sectional cohorts, the ESS measurement was treated as baseline ESS. ESS = Epworth Sleepiness Scale score; CPAP = continuous positive airway pressure.

### Supplementary Figure E2 (Combined Cohort Correlation Plot)

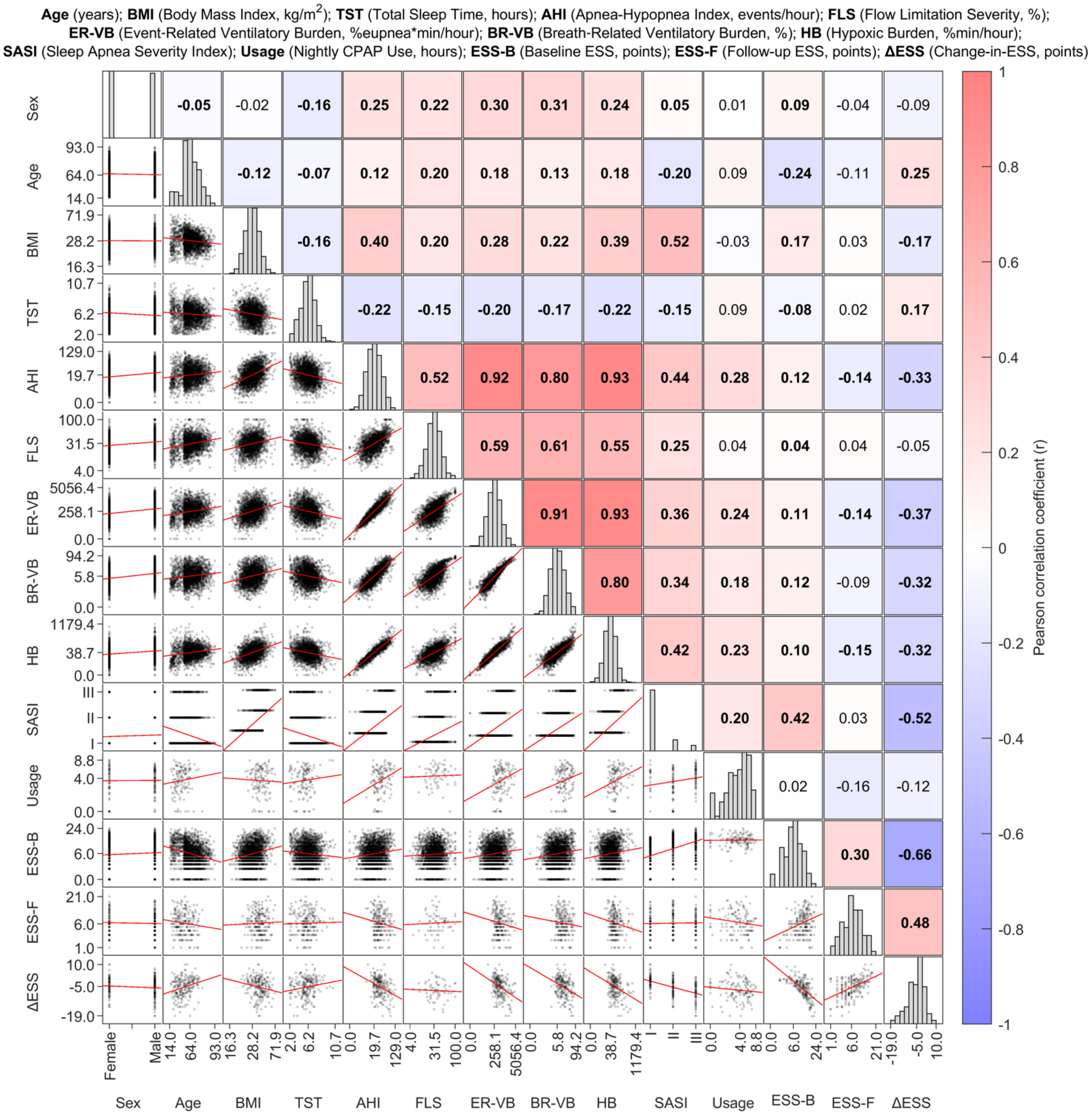

**Supplementary Figure E2.** Histograms and scatter plots of demographic, predictor, and outcome variables (transformed and standardized, with back-transformed tick values) in the combined cohort (maximum $n$ = 2332). The combined cohort consisted of HomePAP home, HomePAP in-lab, BestAIR, ABC, STAGES, and MESA participants. Not all variables were measured in all cohorts, e.g. follow-up ESS was not measured in STAGES and MESA cohorts due to their cross-sectional nature which prevented calculation of change-in-ESS. In cross-sectional cohorts, the single ESS measurement was treated as baseline ESS. Histograms along the diagonal show the distribution of each variable. Scatter plot regression lines are shown in red. Squares are colored and numbered based on Pearson's correlation coefficients (numbers in bold represent $P$ < 0.05). Tick marks represent the smallest, median, and largest values of each variable. Axes scales are non-linear due to transformed data being plotted with back-transformed tick marks. ESS = Epworth Sleepiness Scale score; CPAP = continuous positive airway pressure.

**Supplementary Figure E3 (Statistical Inference in HomePAP Home)**

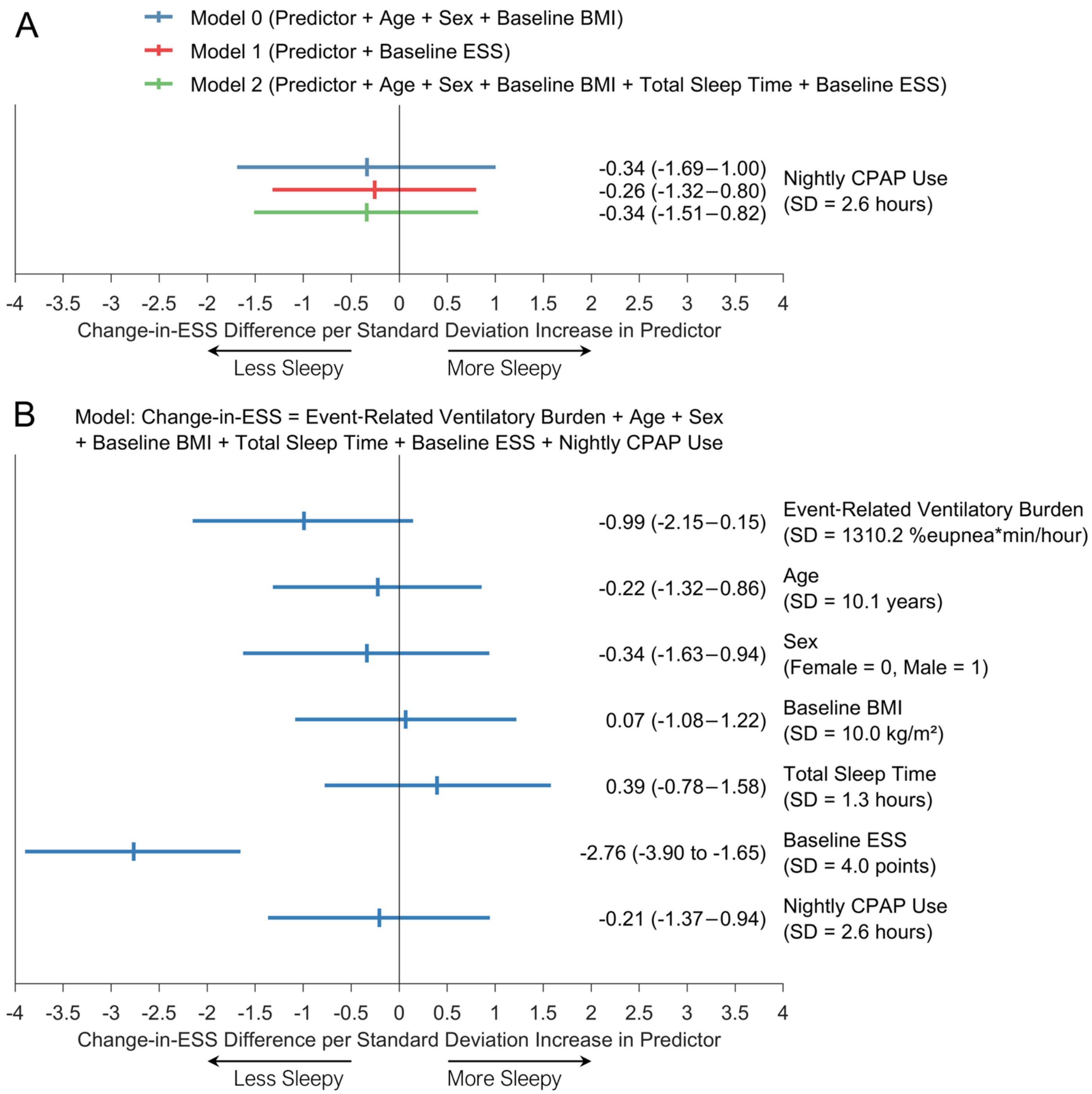

**Supplementary Figure E3.** Association between 1-SD increase in predictor variables and change-in-ESS in the HomePAP home polysomnography cohort. Vertical ticks represent coefficient estimates and horizontal bars represent 95% confidence intervals. (*A*) Linear regression models were adjusted for age, sex, and baseline BMI (model 0, blue bars); adjusted for baseline ESS (model 1, red bars); and further adjusted for age, sex, baseline BMI, and total sleep time (model 2, green bars). (*B*) A linear regression model with event-related ventilatory burden, age, sex, baseline BMI, total sleep time, baseline ESS, and nightly CPAP use. All analyses had *n* = 63 participants because one participant was missing nightly CPAP use data. ESS = Epworth Sleepiness Scale score.

**Supplementary Table E2 (Stepwise Linear Regression)**

**Supplementary Table E2.** Performance of change-in-ESS prediction models using stepwise linear regression selection of variables using data from HomePAP home polysomnograms (*n* = 64). Leave-one-out cross-validation (CV) was performed, and final models were trained in all HomePAP home participants. ESS = Epworth Sleepiness Scale score; $R^2$ = coefficient of determination between predicted change-in-ESS vs actual change-in-ESS; Adj. $R^2$ = adjusted $R^2$; RMSE = root-mean-square error (interpreted as average change-in-ESS error per patient); SASI = Sleep Apnea Severity Index.

**Using conventional and novel parameters (excluding the SASI in the candidate variable list)**

| **Final RMSE** | **Final Adj. $R^2$** | **CV RMSE** | **CV Adj. $R^2$** |
|---|---|---|---|
| 3.88 | 0.322 | 4.32 | 0.163 |
| **Final Variable** | **Coefficient** | **P-Value** | |
| Baseline ESS | -0.883 | < 0.001 | |
| Event-Related Ventilatory Burden | -0.255 | 0.039 | |
| **Cross-Validation Variable** | **Number of Times Selected (Total 64)** | | |
| Age | 0 | | |
| Sex | 0 | | |
| Baseline BMI | 0 | | |
| Total Sleep Time | 0 | | |
| Baseline ESS | 64 | | |
| Apnea-Hypopnea Index | 0 | | |
| Flow Limitation Frequency | 0 | | |
| Flow Limitation Severity | 0 | | |
| Event-Related Ventilatory Burden | 55 | | |
| Breath-Related Ventilatory Burden | 0 | | |
| Desaturation Severity | 0 | | |
| Hypoxic Burden | 0 | | |
| Oxygen Desaturation Index | 0 | | |
| Lowest Oxygen Saturation | 0 | | |

**Using conventional parameters only**

| **Final RMSE** | **Final Adj. $R^2$** | **CV RMSE** | **CV Adj. $R^2$** |
|---|---|---|---|
| 3.99 | 0.296 | 4.23 | 0.205 |
| **Final Variable** | **Coefficient** | **P-Value** | |
| Baseline ESS | -0.943 | < 0.001 | |

| Cross-Validation Variable | Number of Times Selected (Total 64) |
|---|---|
| Age | 0 |
| Sex | 0 |
| Baseline BMI | 0 |
| Total Sleep Time | 0 |
| Baseline ESS | 64 |
| Apnea-Hypopnea Index | 6 |
| Oxygen Desaturation Index | 0 |
| Lowest Oxygen Saturation | 0 |

**Only including the SASI in the candidate variable list**

| Final RMSE | Final Adj. $R^2$ | CV RMSE | CV Adj. $R^2$ |
|---|---|---|---|
| 4.58 | 0.057 | 5.37 | -0.278 |
| **Final Variable** | **Coefficient** | **P-Value** | |
| Sleep Apnea Severity Index (2) | 0.088 | 0.056 | |
| Sleep Apnea Severity Index (1) | 0.115 | 0.039 | |
| **Cross-Validation Variable** | **Number of Times Selected (Total 64)** | | |
| Sleep Apnea Severity Index | 0 | | |
| Sleep Apnea Severity Index (2) | 31 | | |
| Sleep Apnea Severity Index (1) | 31 | | |

**Supplementary Figure E4 (LASSO Selection of Variables)**

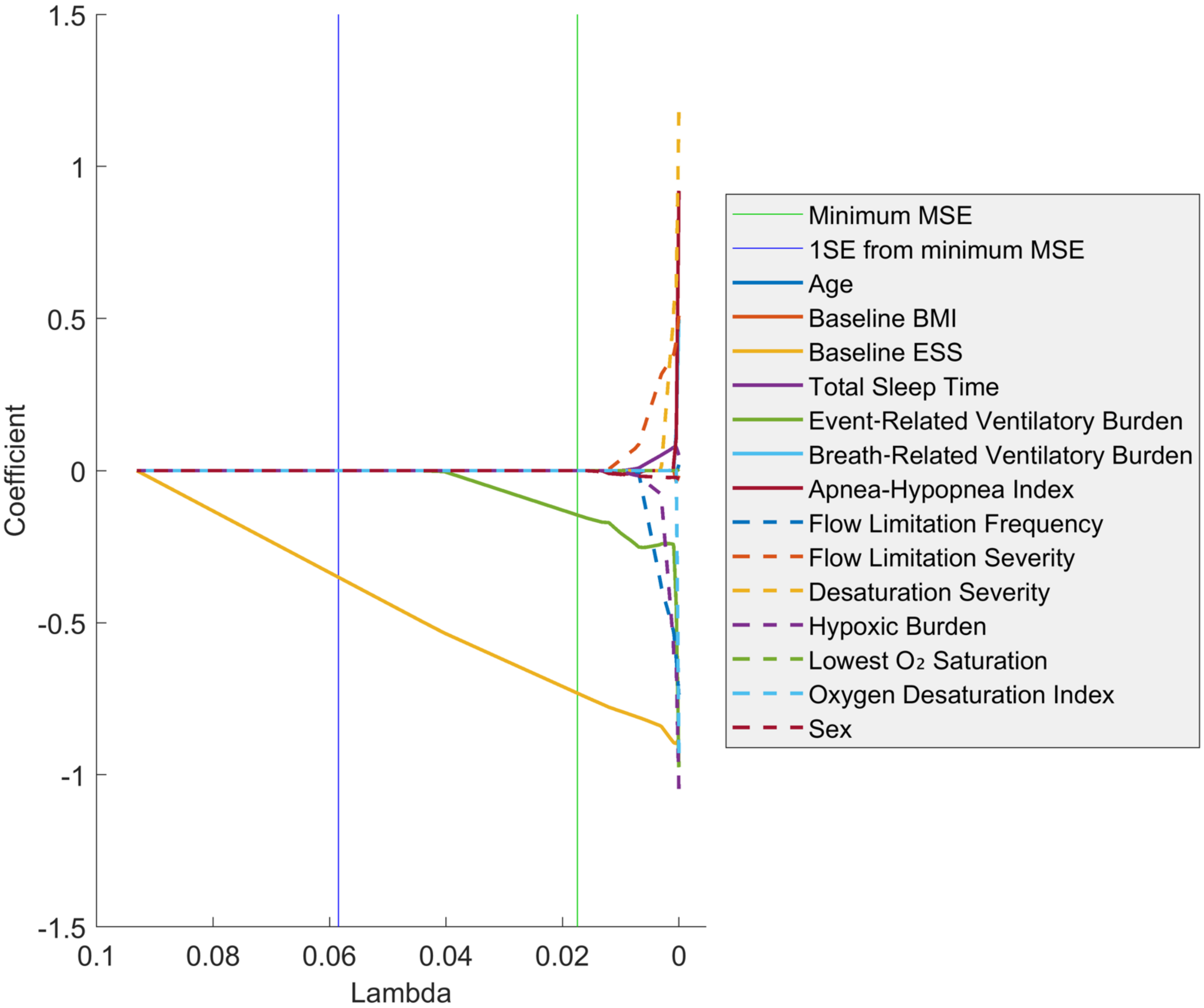

**Supplementary Figure E4.** Linear regression with LASSO selection of baseline variables to predict change-in-ESS using data from HomePAP home polysomnograms (*n* = 64). Variables are selected when their coefficient value deviates from zero. As lambda decreases, the penalty of large coefficient values decreases, which leads to more variables being selected. Note that all variables are eventually selected as lambda approaches zero. Mean squared error (MSE) is a measure of how well the model fits the data, and smaller MSE values indicate better fits. Baseline ESS and event-related ventilatory burden are the first two variables to be selected and are the only selected variables at the minimum MSE line. SE = standard error.

**Supplementary Figure E5 (ROC Curve)**

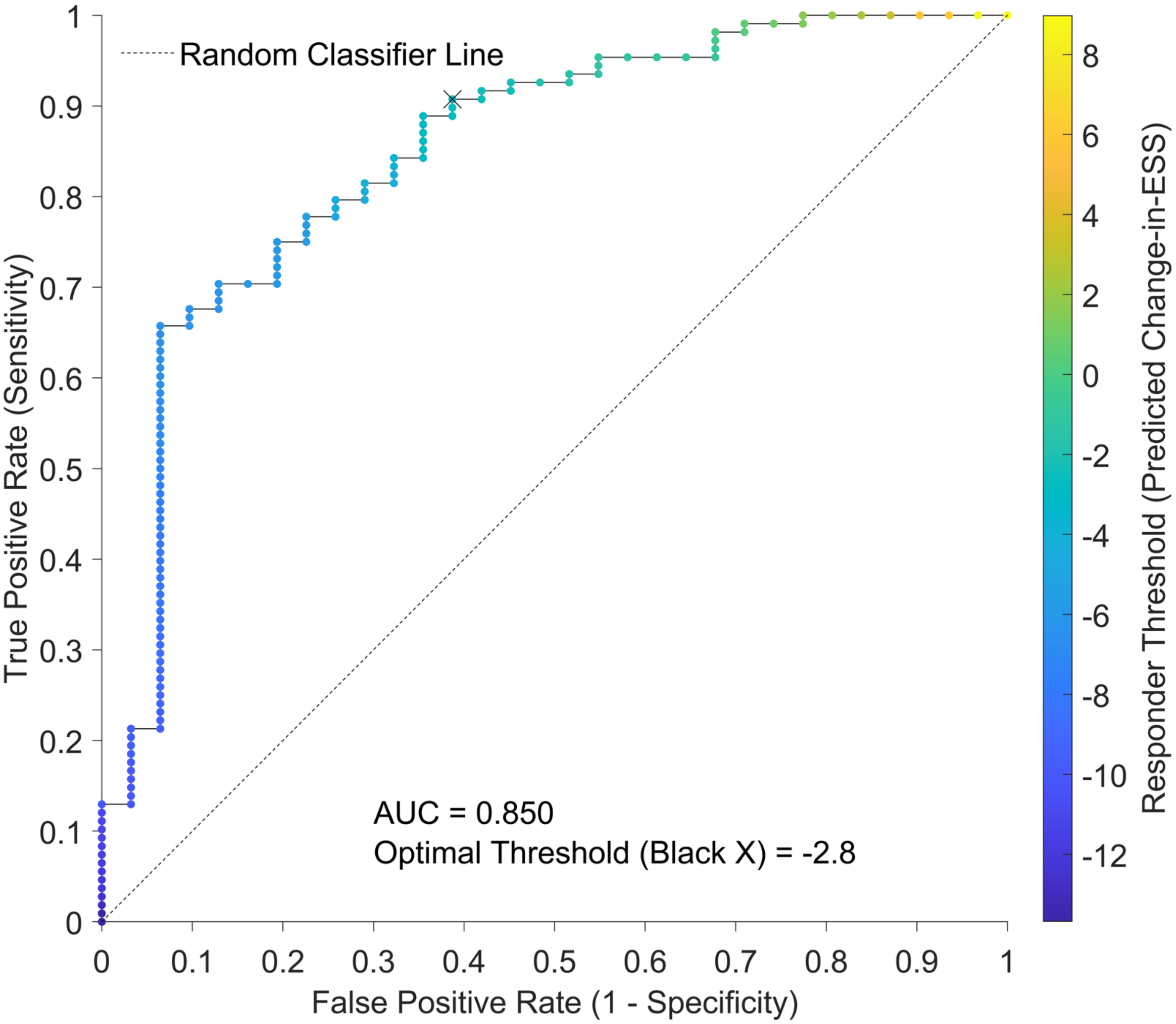

**Supplementary Figure E5.** ROC curve of the linear regression model trained on participants from the training dataset and applying a responder/non-responder change-in-ESS cut-off point of -2, which is the minimum clinically important difference of ESS in obstructive sleep apnea (23, 24). The model was *Change-in-ESS = Baseline ESS + Baseline Event-Related Ventilatory Burden + Intercept*. Data points and color are based on varying the responder threshold. The black cross on the ROC curve represents the optimal probability threshold as determined by the ROC convex hull method (25) with equal false positive and false negative penalties. False positives and false negatives were given a penalty weight of 1, and true positives and true negatives were given a penalty weight of 0. ROC = receiver operating characteristic; AUC = area under the ROC curve.

**Supplementary Table E3 (Classification Sensitivity Analysis)**

**Supplementary Table E3.** Sensitivity analyses of the linear regression model used as a classifier to predict responders and non-responders.

| **Training Dataset** | | | | | | | | | |
|---|---|---|---|---|---|---|---|---|---|
| **Change-in-ESS threshold** | -1 | -2 | -3 | -4 | -5 | -6 | -7 | -8 | -9 |
| **Actual responders (%)** | 82.7 | 77.7 | 70.5 | 63.3 | 56.8 | 49.6 | 43.9 | 38.8 | 30.9 |
| **AUC** | 0.826 | 0.850 | 0.855 | 0.831 | 0.858 | 0.870 | 0.888 | 0.893 | 0.892 |
| **Optimal change-in-ESS threshold** | 0.1 | -2.8 | -2.8 | -2.8 | -6.2 | -6.2 | -7.8 | -7.8 | -7.4 |
| **Accuracy (%)** | 88.5 | 84.2 | 84.2 | 79.9 | 79.9 | 79.9 | 81.3 | 82.0 | 82.7 |
| **Cohen's Kappa** | 0.498 | 0.533 | 0.584 | 0.523 | 0.589 | 0.598 | 0.612 | 0.615 | 0.625 |
| **F1 score** | 0.934 | 0.899 | 0.894 | 0.859 | 0.823 | 0.811 | 0.764 | 0.757 | 0.755 |
| **Sensitivity (%)** | 98.3 | 90.7 | 94.9 | 96.6 | 82.3 | 87.0 | 68.9 | 72.2 | 86.0 |
| **Specificity (%)** | 41.7 | 61.3 | 58.5 | 51.0 | 76.7 | 72.9 | 91.0 | 88.2 | 81.3 |
| **PPV (%)** | 89.0 | 89.1 | 84.5 | 77.3 | 82.3 | 75.9 | 85.7 | 79.6 | 67.3 |
| **NPV (%)** | 83.3 | 65.5 | 82.8 | 89.7 | 76.7 | 85.0 | 78.9 | 83.3 | 92.9 |
| **Holdout Dataset** | | | | | | | | | |
| **Change-in-ESS threshold** | -1 | -2 | -3 | -4 | -5 | -6 | -7 | -8 | -9 |
| **Actual responders (%)** | 71.6 | 63.5 | 50.0 | 45.9 | 39.2 | 31.1 | 27.0 | 25.7 | 23.0 |
| **Accuracy (%)** | 82.4 | 79.7 | 71.6 | 67.6 | 78.4 | 73.0 | 78.4 | 79.7 | 81.1 |
| **Cohen's Kappa** | 0.504 | 0.537 | 0.432 | 0.375 | 0.552 | 0.424 | 0.484 | 0.511 | 0.541 |
| **F1 score** | 0.887 | 0.851 | 0.769 | 0.727 | 0.733 | 0.630 | 0.636 | 0.651 | 0.667 |
| **Sensitivity (%)** | 96.2 | 91.5 | 94.6 | 94.1 | 75.9 | 73.9 | 70.0 | 73.7 | 82.4 |
| **Specificity (%)** | 47.6 | 59.3 | 48.6 | 45.0 | 80.0 | 72.5 | 81.5 | 81.8 | 80.7 |
| **PPV (%)** | 82.3 | 79.6 | 64.8 | 59.3 | 71.0 | 54.8 | 58.3 | 58.3 | 56.0 |
| **NPV (%)** | 83.3 | 80.0 | 90.0 | 90.0 | 83.7 | 86.0 | 88.0 | 90.0 | 93.9 |

The responder/non-responder change-in-ESS cut-off point was varied from -1 to -9. The linear regression model based on the equation *Change-in-ESS = Baseline ESS + Baseline Event-Related Ventilatory Burden + Intercept* was developed in the training dataset and the optimal threshold was determined using the convex hull method with equal false positive and false negative penalties. The model and optimal threshold were applied to the training and holdout datasets to determine accuracy, Cohen's kappa, F1 score, sensitivity, specificity, PPV, and NPV. AUC = area under the receiver operator curve; PPV = positive predictive value; NPV = negative predictive value.

**Supplementary Table E4 (Alternative Classification Models)**

**Supplementary Table E4.** Performance of linear regression models used for classification in the training and holdout datasets. The change-in-ESS cut-point used to differentiate responders from non-responders was -2, which is the minimum clinically important difference of ESS in obstructive sleep apnea (23, 24). Each model was developed in the training dataset and the optimal responder threshold was found using the convex hull method with equal false positive and false negative penalties. The model and optimal threshold were applied in the holdout dataset. AUC = area under the receiver operator curve; PPV = positive predictive value; NPV = negative predictive value.

**Training Dataset**

| Baseline Predictors | AUC | Optimal Threshold | Accuracy (%) | Cohen's Kappa | F1 Score | Sensitivity (%) | Specificity (%) | PPV (%) | NPV (%) |
|---|---|---|---|---|---|---|---|---|---|
| ESS | 0.831 | -0.4 | 83.5 | 0.389 | 0.902 | 98.1 | 32.3 | 83.5 | 83.3 |
| Sleep Apnea Severity Index | 0.755 | -1.7 | 77.7 | 0.000 | 0.874 | 100.0 | 0.0 | 77.7 | – |
| ESS + Apnea-Hypopnea Index | 0.850 | -2.0 | 85.6 | 0.554 | 0.910 | 93.5 | 58.1 | 88.6 | 72.0 |
| ESS + Event-Related Ventilatory Burden | 0.850 | -2.8 | 84.2 | 0.533 | 0.899 | 90.7 | 61.3 | 89.1 | 65.5 |
| ESS + Breath-Related Ventilatory Burden | 0.843 | -2.4 | 84.9 | 0.526 | 0.906 | 93.5 | 54.8 | 87.8 | 70.8 |
| ESS + Desaturation Severity | 0.844 | -1.9 | 84.2 | 0.497 | 0.902 | 93.5 | 51.6 | 87.1 | 69.6 |
| ESS + Hypoxic Burden | 0.849 | -2.2 | 84.9 | 0.537 | 0.905 | 92.6 | 58.1 | 88.5 | 69.2 |
| ESS + Oxygen Desaturation Index | 0.845 | -2.5 | 84.2 | 0.533 | 0.899 | 90.7 | 61.3 | 89.1 | 65.5 |
| ESS + Lowest Oxygen Saturation | 0.832 | -1.6 | 84.2 | 0.484 | 0.903 | 94.4 | 48.4 | 86.4 | 71.4 |

**Holdout Dataset**

| Baseline Predictors | Accuracy (%) | Cohen's Kappa | F1 Score | Sensitivity (%) | Specificity (%) | PPV (%) | NPV (%) |
|---|---|---|---|---|---|---|---|
| ESS | 77.0 | 0.438 | 0.844 | 97.9 | 40.7 | 74.2 | 91.7 |
| Sleep Apnea Severity Index | 62.2 | -0.027 | 0.767 | 97.9 | 0.0 | 63.0 | 0.0 |
| ESS + Apnea-Hypopnea Index | 77.0 | 0.457 | 0.838 | 93.6 | 48.1 | 75.9 | 81.2 |
| ESS + Event-Related Ventilatory Burden | 79.7 | 0.537 | 0.851 | 91.5 | 59.3 | 79.6 | 80.0 |
| ESS + Breath-Related Ventilatory Burden | 78.4 | 0.494 | 0.846 | 93.6 | 51.9 | 77.2 | 82.4 |
| ESS + Desaturation Severity | 75.7 | 0.420 | 0.830 | 93.6 | 44.4 | 74.6 | 80.0 |
| ESS + Hypoxic Burden | 77.0 | 0.457 | 0.838 | 93.6 | 48.1 | 75.9 | 81.2 |
| ESS + Oxygen Desaturation Index | 78.4 | 0.502 | 0.843 | 91.5 | 55.6 | 78.2 | 78.9 |
| ESS + Lowest Oxygen Saturation | 78.4 | 0.476 | 0.852 | 97.9 | 44.4 | 75.4 | 92.3 |

**Supplementary Results 1 (Classification Performance Using Smaller False Positive Penalty)**

ROC Curve Using Smaller False Positive Penalty (Supplementary Figure E6)

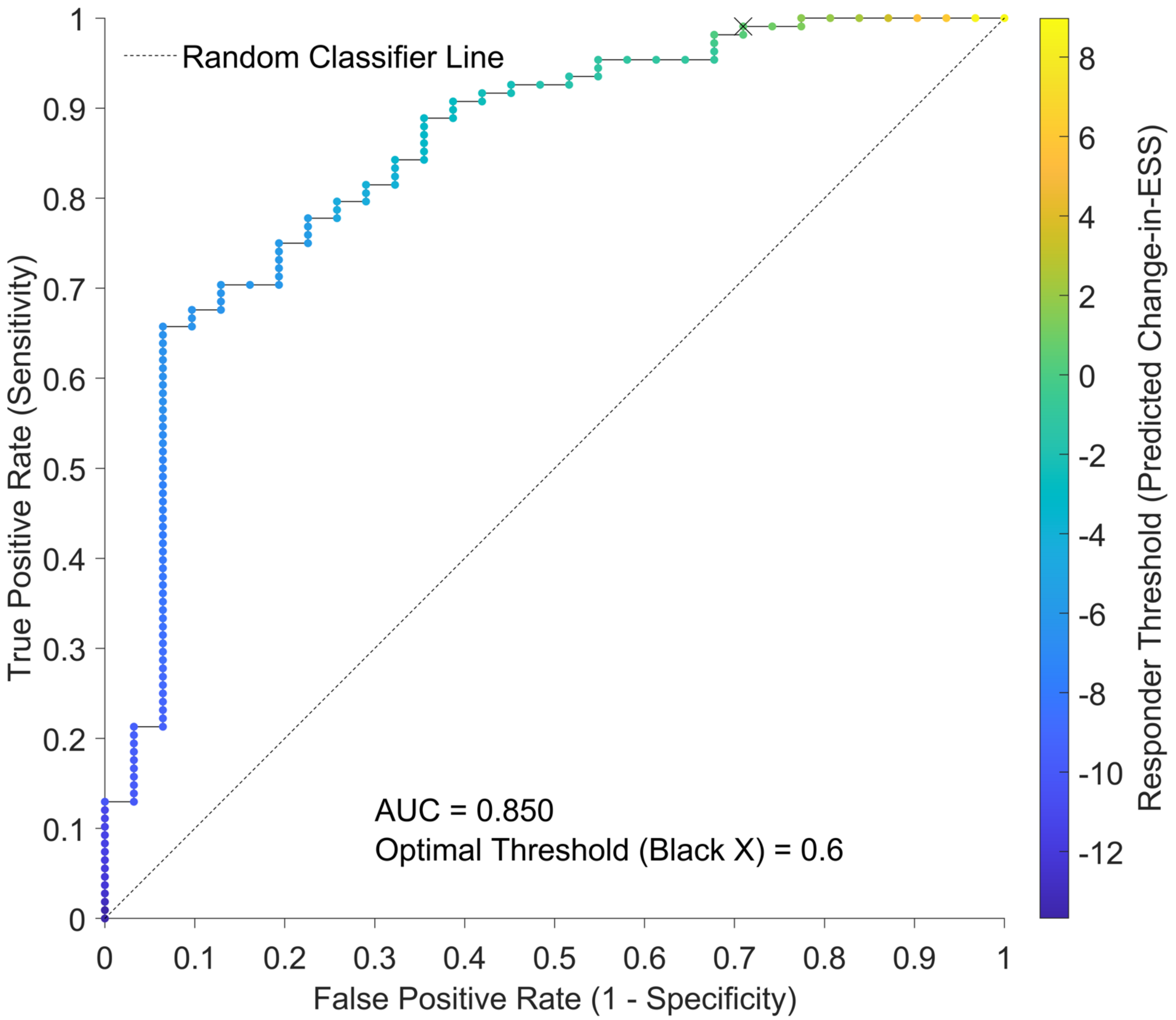

**Supplementary Figure E6.** ROC curve of the linear regression model trained on participants from the training dataset and applying a responder/non-responder change-in-ESS cut-off point of -2, which is the minimum clinically important difference of ESS in obstructive sleep apnea (23, 24). The model was *Change-in-ESS = Baseline ESS + Baseline Event-Related Ventilatory Burden + Intercept*. Data points and color are based on varying the responder threshold. The black cross on the ROC curve represents the optimal threshold as determined by the ROC convex hull method (25) with smaller false positive penalty. False positives were given a penalty weight of 0.5, false negatives were given a penalty weight of 1, and true positives and true negatives were given a penalty weight of 0. ROC = receiver operating characteristic; AUC = area under the ROC curve.

Classification Performance Using Smaller False Positive Penalty (Figure E7)

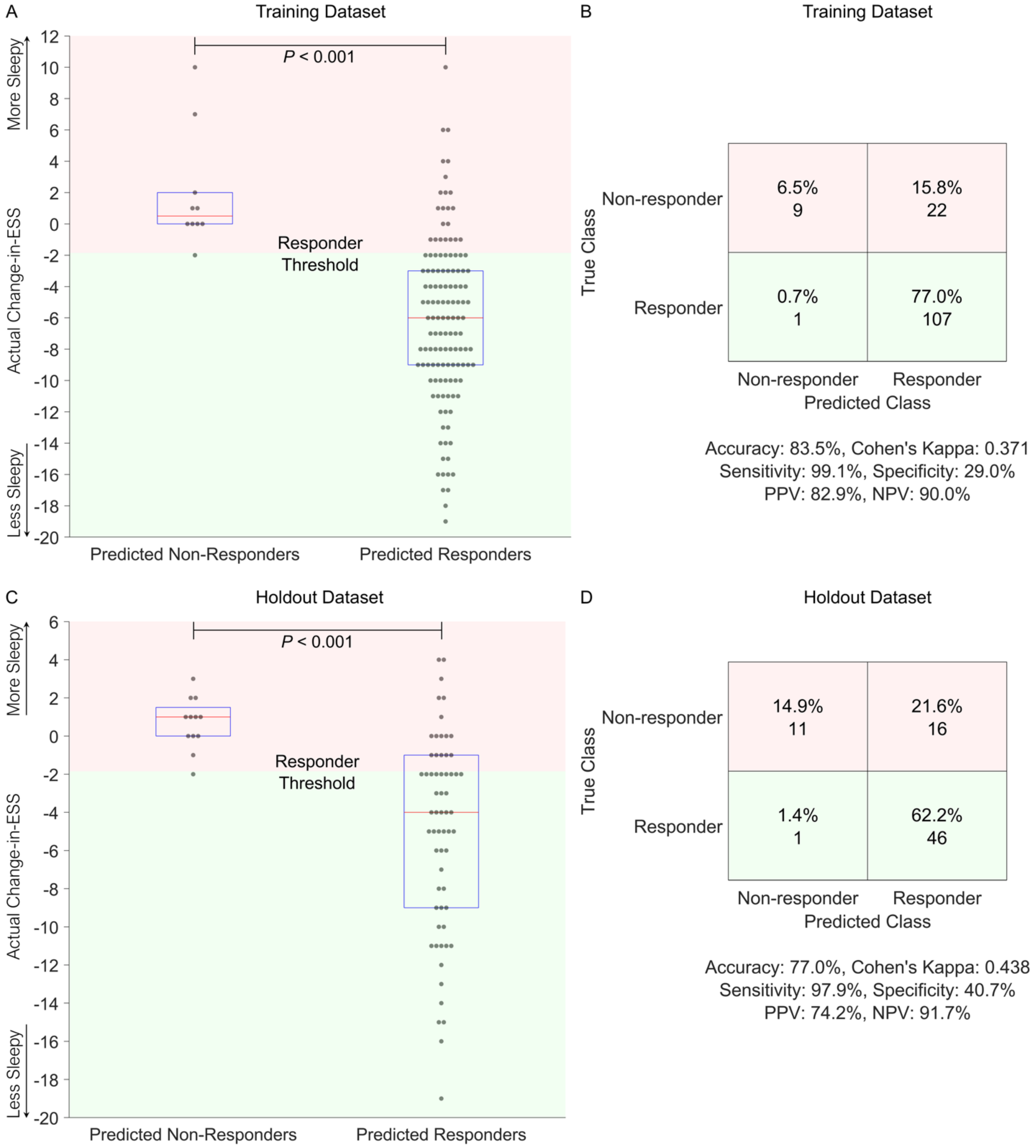

**Supplementary Figure E7.** Boxplots and confusion matrices of results using the linear regression model as a classifier in the training and holdout datasets. (*A*) and (*C*) are boxplots representing the 25$^{th}$ percentile (lower horizontal line), median (red line), and 75$^{th}$ percentile (upper horizontal line). Each participant is represented as a single point and random horizontal jitter was added. The Mann-Whitney U-test was performed between predicted responder and non-responder groups. (*B*) and (*D*) are confusion matrices with summary performance metrics presented underneath. The change-in-ESS cut-point used to differentiate responders from non-responders was -2, which is the minimum clinically important difference of ESS in obstructive sleep apnea (23, 24). The model *Change-in-ESS = Baseline ESS + Baseline Event-Related Ventilatory Burden + Intercept* was developed in the training dataset and the optimal responder threshold was determined by the ROC convex hull method (25) with smaller false positive penalty. False positives were given a penalty weight of 0.5, false negatives were given a penalty weight of 1, and true positives and true negatives were given a penalty weight of 0. The model and optimal threshold were applied in the holdout dataset. PPV = positive predictive value; NPV = negative predictive value.

Classification Sensitivity Analysis Using Smaller False Positive Penalty (Supplementary Table E5)

**Supplementary Table E5.** Sensitivity analyses of the linear regression model used as a classifier to predict responders and non-responders.

| **Training Dataset** | | | | | | | | | |
|---|---|---|---|---|---|---|---|---|---|
| **Change-in-ESS threshold** | -1 | -2 | -3 | -4 | -5 | -6 | -7 | -8 | -9 |
| **Actual responders (%)** | 82.7 | 77.7 | 70.5 | 63.3 | 56.8 | 49.6 | 43.9 | 38.8 | 30.9 |
| **AUC** | 0.826 | 0.850 | 0.855 | 0.831 | 0.858 | 0.870 | 0.888 | 0.893 | 0.892 |
| **Optimal change-in-ESS threshold** | 0.6 | 0.6 | -2.8 | -2.8 | -2.8 | -6.2 | -6.2 | -6.2 | -7.4 |
| **Accuracy (%)** | 88.5 | 83.5 | 84.2 | 79.9 | 76.3 | 79.9 | 79.9 | 79.1 | 82.7 |
| **Cohen's Kappa** | 0.476 | 0.371 | 0.584 | 0.523 | 0.484 | 0.598 | 0.604 | 0.595 | 0.625 |
| **F1 score** | 0.934 | 0.903 | 0.894 | 0.859 | 0.825 | 0.811 | 0.800 | 0.782 | 0.755 |
| **Sensitivity (%)** | 99.1 | 99.1 | 94.9 | 96.6 | 98.7 | 87.0 | 91.8 | 96.3 | 86.0 |
| **Specificity (%)** | 37.5 | 29.0 | 58.5 | 51.0 | 46.7 | 72.9 | 70.5 | 68.2 | 81.3 |
| **PPV (%)** | 88.4 | 82.9 | 84.5 | 77.3 | 70.9 | 75.9 | 70.9 | 65.8 | 67.3 |
| **NPV (%)** | 90.0 | 90.0 | 82.8 | 89.7 | 96.6 | 85.0 | 91.7 | 96.7 | 92.9 |
| **Holdout Dataset** | | | | | | | | | |
| **Change-in-ESS threshold** | -1 | -2 | -3 | -4 | -5 | -6 | -7 | -8 | -9 |
| **Actual responders (%)** | 71.6 | 63.5 | 50.0 | 45.9 | 39.2 | 31.1 | 27.0 | 25.7 | 23.0 |
| **Accuracy (%)** | 82.4 | 77.0 | 71.6 | 67.6 | 66.2 | 73.0 | 74.3 | 75.7 | 81.1 |
| **Cohen's Kappa** | 0.504 | 0.438 | 0.432 | 0.375 | 0.385 | 0.424 | 0.445 | 0.472 | 0.541 |
| **F1 score** | 0.887 | 0.844 | 0.769 | 0.727 | 0.699 | 0.630 | 0.627 | 0.640 | 0.667 |
| **Sensitivity (%)** | 96.2 | 97.9 | 94.6 | 94.1 | 100.0 | 73.9 | 80.0 | 84.2 | 82.4 |
| **Specificity (%)** | 47.6 | 40.7 | 48.6 | 45.0 | 44.4 | 72.5 | 72.2 | 72.7 | 80.7 |
| **PPV (%)** | 82.3 | 74.2 | 64.8 | 59.3 | 53.7 | 54.8 | 51.6 | 51.6 | 56.0 |
| **NPV (%)** | 83.3 | 91.7 | 90.0 | 90.0 | 100.0 | 86.0 | 90.7 | 93.0 | 93.9 |

The responder/non-responder change-in-ESS cut-off point was varied from -1 to -9. The linear regression model based on the equation *Change-in-ESS = Baseline ESS + Baseline Event-Related Ventilatory Burden + Intercept* was developed in the training dataset and the optimal threshold was determined using the convex hull method with smaller false positive penalty. The model and optimal threshold were applied to the training and holdout datasets to determine accuracy, Cohen's kappa, F1 score, sensitivity, specificity, PPV, and NPV. AUC = area under the receiver operator curve; PPV = positive predictive value; NPV = negative predictive value.

Alternative Classification Models Using Smaller False Positive Penalty (Supplementary Table E6)

**Supplementary Table E6.** Performance of linear regression models used for classification in the training and holdout datasets. The change-in-ESS cut-point used to differentiate responders from non-responders was -2, which is the minimum clinically important difference of ESS in obstructive sleep apnea (23, 24). Each model was developed in the training dataset and the optimal responder threshold was found using the convex hull method with smaller false positive penalty. The model and optimal threshold were applied in the holdout dataset. AUC = area under the receiver operator curve; PPV = positive predictive value; NPV = negative predictive value.

**Training Dataset**

| Baseline Predictors | AUC | Optimal Threshold | Accuracy (%) | Cohen's Kappa | F1 Score | Sensitivity (%) | Specificity (%) | PPV (%) | NPV (%) |
|---|---|---|---|---|---|---|---|---|---|
| ESS | 0.831 | -0.4 | 83.5 | 0.389 | 0.902 | 98.1 | 32.3 | 83.5 | 83.3 |
| Sleep Apnea Severity Index | 0.755 | -1.7 | 77.7 | 0.000 | 0.874 | 100.0 | 0.0 | 77.7 | – |
| ESS + Apnea-Hypopnea Index | 0.850 | 0.4 | 83.5 | 0.371 | 0.903 | 99.1 | 29.0 | 82.9 | 90.0 |
| ESS + Event-Related Ventilatory Burden | 0.850 | 0.6 | 83.5 | 0.371 | 0.903 | 99.1 | 29.0 | 82.9 | 90.0 |
| ESS + Breath-Related Ventilatory Burden | 0.843 | 1.5 | 83.5 | 0.351 | 0.904 | 100.0 | 25.8 | 82.4 | 100.0 |
| ESS + Desaturation Severity | 0.844 | 0.6 | 84.2 | 0.407 | 0.907 | 99.1 | 32.3 | 83.6 | 90.9 |
| ESS + Hypoxic Burden | 0.849 | 0.2 | 84.2 | 0.407 | 0.907 | 99.1 | 32.3 | 83.6 | 90.9 |
| ESS + Oxygen Desaturation Index | 0.845 | 0.7 | 83.5 | 0.371 | 0.903 | 99.1 | 29.0 | 82.9 | 90.0 |
| ESS + Lowest Oxygen Saturation | 0.832 | 2.2 | 82.7 | 0.312 | 0.900 | 100.0 | 22.6 | 81.8 | 100.0 |

**Holdout Dataset**

| Baseline Predictors | Accuracy (%) | Cohen's Kappa | F1 Score | Sensitivity (%) | Specificity (%) | PPV (%) | NPV (%) |
|---|---|---|---|---|---|---|---|
| ESS | 77.0 | 0.438 | 0.844 | 97.9 | 40.7 | 74.2 | 91.7 |
| Sleep Apnea Severity Index | 62.2 | -0.027 | 0.767 | 97.9 | 0.0 | 63.0 | 0.0 |
| ESS + Apnea-Hypopnea Index | 77.0 | 0.438 | 0.844 | 97.9 | 40.7 | 74.2 | 91.7 |
| ESS + Event-Related Ventilatory Burden | 77.0 | 0.438 | 0.844 | 97.9 | 40.7 | 74.2 | 91.7 |
| ESS + Breath-Related Ventilatory Burden | 73.0 | 0.320 | 0.821 | 97.9 | 29.6 | 70.8 | 88.9 |
| ESS + Desaturation Severity | 75.7 | 0.399 | 0.836 | 97.9 | 37.0 | 73.0 | 90.9 |
| ESS + Hypoxic Burden | 77.0 | 0.438 | 0.844 | 97.9 | 40.7 | 74.2 | 91.7 |
| ESS + Oxygen Desaturation Index | 77.0 | 0.438 | 0.844 | 97.9 | 40.7 | 74.2 | 91.7 |
| ESS + Lowest Oxygen Saturation | 68.9 | 0.196 | 0.800 | 97.9 | 18.5 | 67.6 | 83.3 |

**Supplementary Results 2 (%Change-in-ESS as the Outcome Variable)**

Percent change-in-ESS (%change-in-ESS) has been used as an outcome measure in at least one study (27). In secondary exploratory analyses, %change-in-ESS was used as the outcome measure instead of change-in-ESS. Associations between OSA severity measures and %change-in-ESS were similar, except that hypoxic burden had the highest effect size instead of event-related ventilatory burden (Figure E8). This is reasonable given the strong correlation between hypoxic burden and event-related ventilatory burden (Pearson's $r$=0.93, Figure E2). However, %change-in-ESS could not be predicted using stepwise linear regression and LASSO selection of variables. Using stepwise linear regression, only baseline ESS was chosen and the adjusted $R^2$ between predicted change-in-ESS and actual change-in-ESS was -0.008 (Table E7). Using LASSO selection of variables, baseline ESS and hypoxic burden were selected (Figure E9). We posit that %change-in-ESS does not represent true sleepiness change. For example, a patient whose ESS decreases from 20 to 10 has a 50% change-in-ESS. A patient whose ESS decreases from 2 to 1 also has a 50% change-in-ESS. However, the second patient's ESS improvement may not be modulated by their baseline OSA severity or ESS.

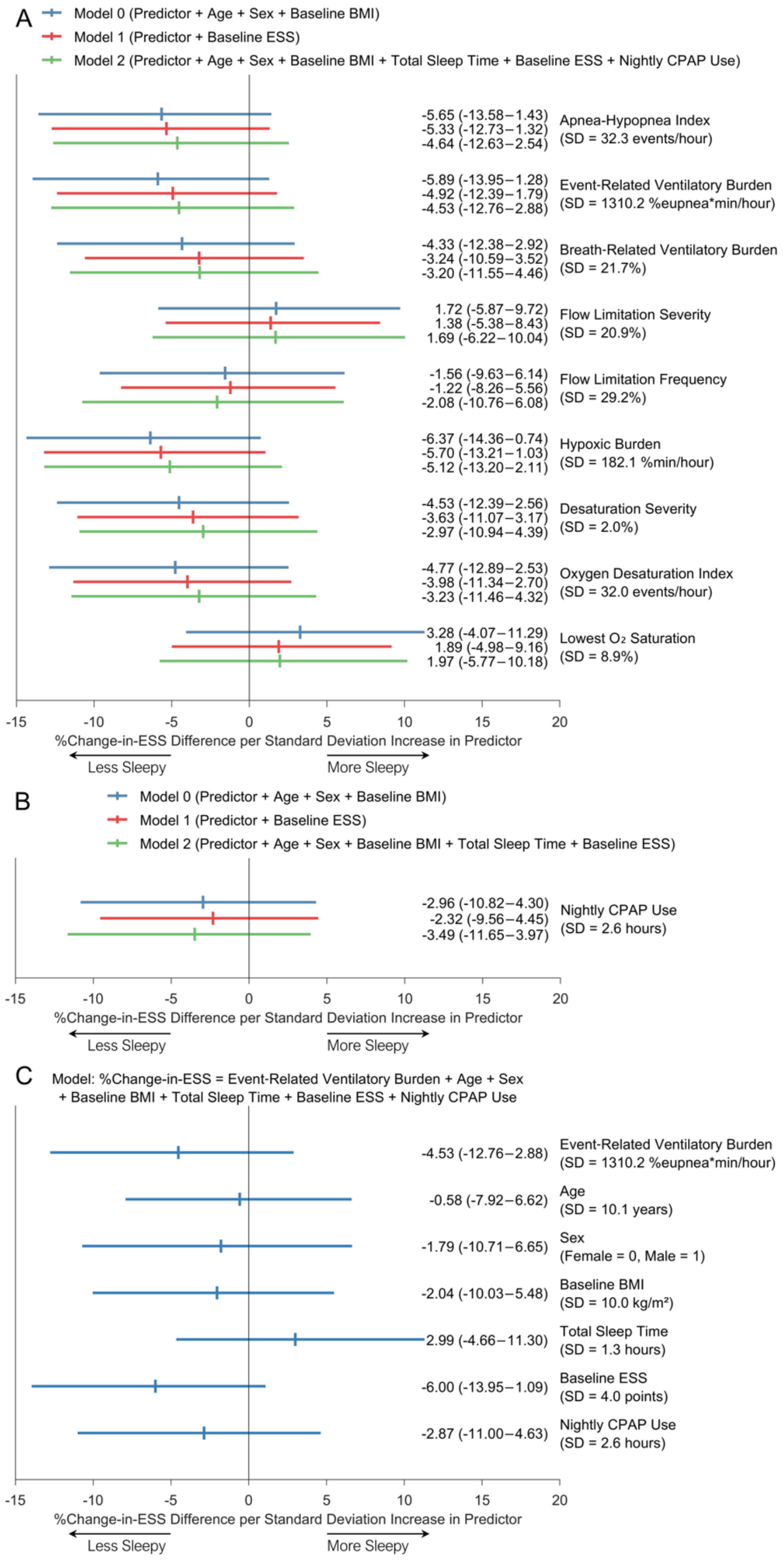

**Supplementary Figure E8.** Association between 1-SD increase in baseline predictor variables and %change-in-ESS in the HomePAP home polysomnography cohort. Vertical ticks represent coefficient estimates and horizontal bars represent 95% confidence intervals. (*A*) Linear regression models were adjusted for age, sex, and baseline BMI (model 0, blue bars); adjusted for baseline ESS (model 1, red bars); and further adjusted for age, sex, baseline BMI, total sleep time, and nightly CPAP use (model 2, green bars). (*B*) Linear regression models were adjusted identically to (*A*), except model 2 which was not adjusted for CPAP use. (*C*) Linear regression model with event-related ventilatory burden, age, sex, baseline BMI, total sleep time, baseline ESS, and nightly CPAP use. All analyses had *n* = 64 participants except for those with nightly CPAP use, which had *n* = 63 due to one participant missing this data. ESS = Epworth Sleepiness Scale score.

**Supplementary Table E7.** Performance of %change-in-ESS prediction models using stepwise linear regression selection of variables using data from HomePAP home polysomnograms (*n* = 64). Leave-one-out cross-validation (CV) was performed, and final models were trained in all HomePAP home participants. ESS = Epworth Sleepiness Scale score; $R^2$ = coefficient of determination between predicted change-in-ESS vs actual change-in-ESS; Adj. $R^2$ = adjusted $R^2$; RMSE = root-mean-square error (interpreted as average change-in-ESS error per patient); SASI = Sleep Apnea Severity Index.

**Using conventional and novel parameters (excluding the SASI in the candidate variable list)**

| **Final RMSE** | **Final Adj. $R^2$** | **CV RMSE** | **CV Adj. $R^2$** |
|---|---|---|---|
| 27.59 | -0.008 | 31.57 | -0.313 |
| **Final Variable** | **Coefficient** | **P-Value** | |
| Baseline ESS | -0.339 | 0.048 | |
| **Cross-Validation Variable** | **Number of Times Selected (Total 64)** | | |
| Age | 0 | | |
| Sex | 0 | | |
| Baseline BMI | 0 | | |
| Total Sleep Time | 0 | | |
| Baseline ESS | 27 | | |
| Apnea-Hypopnea Index | 1 | | |
| Flow Limitation Frequency | 0 | | |
| Flow Limitation Severity | 0 | | |
| Event-Related Ventilatory Burden | 0 | | |
| Breath-Related Ventilatory Burden | 0 | | |
| Desaturation Severity | 0 | | |
| Hypoxic Burden | 13 | | |
| Oxygen Desaturation Index | 0 | | |
| Lowest Oxygen Saturation | 0 | | |

**Using conventional parameters only**

| **Final RMSE** | **Final Adj. $R^2$** | **CV RMSE** | **CV Adj. $R^2$** |
|---|---|---|---|
| 27.59 | -0.008 | 30.57 | -0.227 |
| **Final Variable** | **Coefficient** | **P-Value** | |
| Baseline ESS | -0.339 | 0.048 | |
| **Cross-Validation Variable** | **Number of Times Selected (Total 64)** | | |
| Age | 0 | | |
| Sex | 0 | | |
| Baseline BMI | 0 | | |

| | |
|---|---|
| Total Sleep Time | 0 |
| Baseline ESS | 30 |
| Apnea-Hypopnea Index | 1 |
| Oxygen Desaturation Index | 0 |
| Lowest Oxygen Saturation | 0 |

**Only including the SASI in the candidate variable list**

| **Final RMSE** | **Final Adj. $R^2$** | **CV RMSE** | **CV Adj. $R^2$** |
|---|---|---|---|
| 28.33 | -0.046 | 28.72 | -0.075 |
| **Cross-Validation Variable** | **Number of Times Selected (Total 64)** | | |
| Sleep Apnea Severity Index | 0 | | |

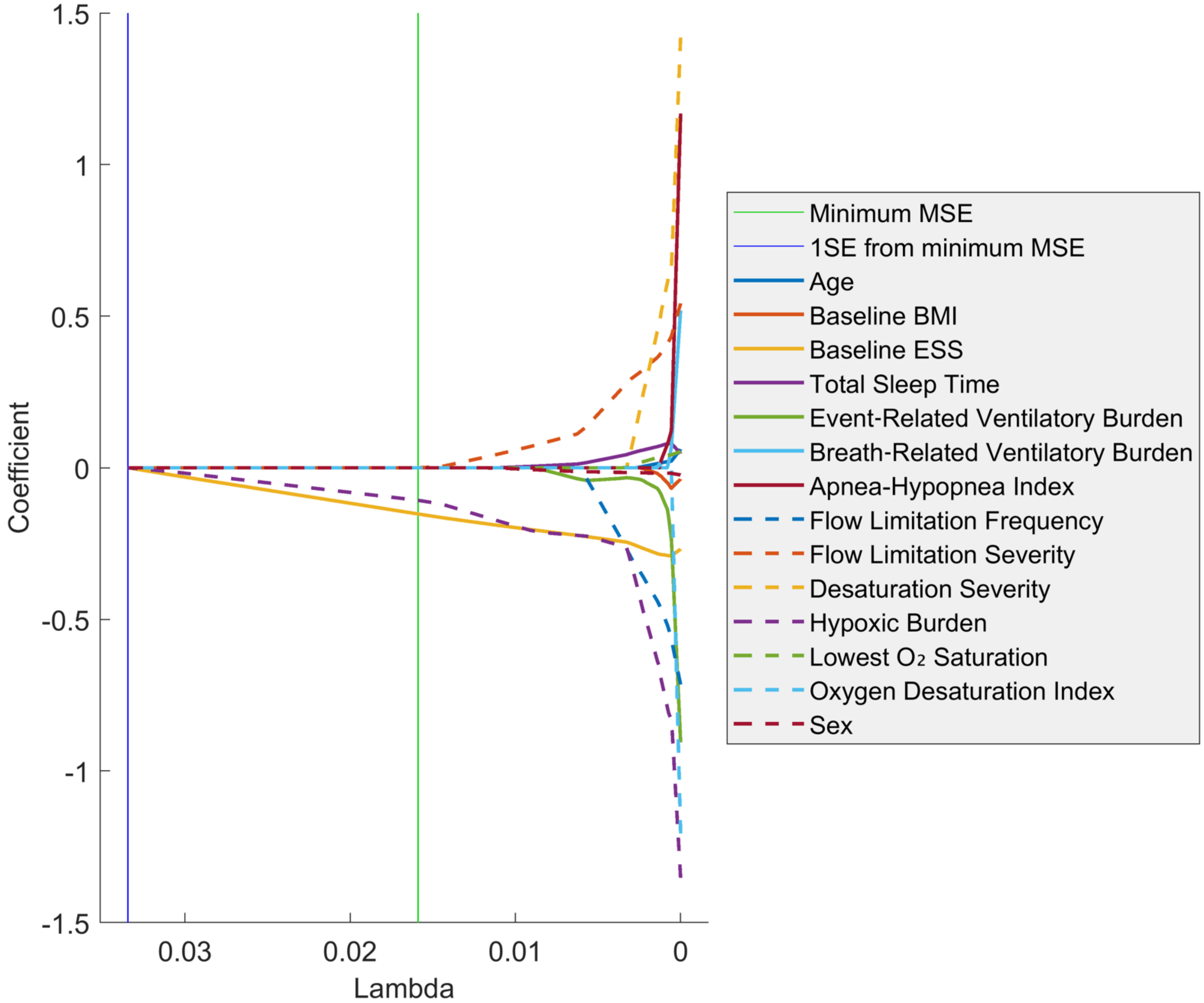

**Supplementary Figure E9.** Linear regression with LASSO selection of baseline variables to predict %change-in-ESS using data from HomePAP home polysomnograms (*n* = 64). Variables are selected when their coefficient value deviates from zero. As lambda decreases, the penalty of large coefficient values decreases, which leads to more variables being selected. Note that all variables are eventually selected as lambda approaches zero. Mean squared error (MSE) is a measure of how well the model fits the data, and smaller MSE values indicate better fits. Baseline ESS and hypoxic burden are the first two variables to be selected and are the only selected variables at the minimum MSE line. SE = standard error.

21. Ribeiro Jr PJ, Diggle PJ. Analysis of Geostatistical Data. The geoR package; 2022.

22. R Core Team R. R: A language and environment for statistical computing. R Foundation for Statistical Computing, Vienna, Austria; 2022.
